# Impact of maternal antibodies and microbiota development on the immunogenicity of oral rotavirus vaccine in African, Indian, and European infants: a prospective cohort study

**DOI:** 10.1101/2020.11.02.20224576

**Authors:** Edward P. K. Parker, Christina Bronowski, Kulandaipalayam Natarajan C. Sindhu, Sudhir Babji, Blossom Benny, Noelia Carmona-Vicente, Nedson Chasweka, End Chinyama, Nigel A. Cunliffe, Queen Dube, Sidhartha Giri, Nicholas C. Grassly, Annai Gunasekaran, Deborah Howarth, Sushil Immanuel, Khuzwayo C. Jere, Beate Kampmann, Jenna Lowe, Jonathan Mandolo, Ira Praharaj, Bakthavatsalam Sandya Rani, Sophia Silas, Vivek Kumar Srinivasan, Mark Turner, Srinivasan Venugopal, Valsan Philip Verghese, Alistair C. Darby, Gagandeep Kang, Miren Iturriza-Gómara

**Affiliations:** The Vaccine Centre, Department of Clinical Research, London School of Hygiene and Tropical Medicine, London, WC1E 7HT, UK; Institute of Infection and Global Health, University of Liverpool, Liverpool, L69 7BE, UK; Division of Gastrointestinal Sciences, Christian Medical College, Vellore, Tamil Nadu, 632004, India; Malawi-Liverpool-Wellcome Trust Clinical Research Programme, University of Malawi, Blantyre, PO Box 30096, Malawi; Department of Medical Laboratory Sciences, College of Medicine, University of Malawi, Private Bag 360, Chichiri, Blantyre 3, Malawi; Department of Infectious Disease Epidemiology, Imperial College London, London, W2 1PG, UK; Institute of Translational Medicine, University of Liverpool, Liverpool, L8 7SS, UK; Department of Child Health, Christian Medical College, Vellore, Tamil Nadu, 632004, India; Institute of Integrative Biology, University of Liverpool, Liverpool, L69 7ZB, UK

**Keywords:** Oral vaccines, environmental enteric dysfunction, intestinal microbiota, maternal antibodies, rotavirus, vaccination

## Abstract

Identifying risk factors for impaired oral rotavirus vaccine (ORV) efficacy in low-income countries may lead to improvements in vaccine design and delivery. We measured maternal rotavirus antibodies, environmental enteric dysfunction (EED), and bacterial gut microbiota development among infants receiving two doses of Rotarix in India (n = 307), Malawi (n = 119), and the UK (n = 60), using standardised methods across cohorts. ORV shedding and seroconversion rates were significantly lower in Malawi and India than the UK. Maternal rotavirus-specific antibodies in serum and breastmilk were negatively correlated with ORV response in India and Malawi, and this was mediated partly by a reduction in ORV replication. In the UK, ORV replication was not inhibited despite comparable maternal antibody levels. In both India and Malawi, pre-vaccination microbiota diversity was negatively correlated with ORV immunogenicity, suggesting that high early-life microbial exposure may contribute to impaired vaccine efficacy.

## Introduction

The roll-out of oral rotavirus vaccine (ORV) has now reached over 100 countries. This initiative – in parallel with advances in sanitation infrastructure and increased use of oral rehydration therapy – has led to substantial declines in global diarrhoeal mortality^1^. Whereas rotavirus was linked with over 500,000 infant deaths annually at the turn of the century^2^, this number currently stands at approximately 130,000^3^. Yet the potential impact of ORV is constrained by the impaired performance of current vaccines in low- and middle-income countries (LMICs). The 1-year protective efficacy of Rotarix against severe rotavirus-associated gastroenteritis is >95% in Europe^4^ but may fall below 50% in sub-Saharan Africa^5^. Moreover, while a variety of interventions to boost ORV performance have been tested (e.g. temporary withholding of breastfeeding), these have generally proven either ineffective or of modest benefit^6^, highlighting the need for new strategies informed by a deeper understanding of the mechanisms underlying the vaccine efficacy gap.

A variety of risk factors have been linked with impaired oral vaccine performance in LMICs^7^. Passively acquired maternal antibodies appear to interfere with ORV immunogenicity ^8^, as documented for a variety of parenteral vaccines ^9^. However, it remains unclear whether rotavirus-specific antibody concentrations and their inhibitory effect are higher in LMICs. Environmental enteric dysfunction (EED) – a subclinical condition associated with intestinal inflammation and increased permeability – is common in LMICs and has been cited as a possible cause of oral vaccine failure^10^. However, EED is generally measured using faecal or plasma biomarkers as a proxy for mucosal immune status, and studies of these biomarkers at the time of oral vaccine delivery have yielded mixed results^11^. The bacterial gut microbiota shapes and is in turn shaped by the developing infant immune system. Among infants in Ghana, Rotarix immunogenicity was positively correlated with the relative abundance of *Streptococcus bovis* and negatively correlated with Bacteroidetes abundance at the time of the first vaccine dose^12^. However, no significant discrepancies in microbiota composition were apparent when comparing Rotarix responders with non-responders in India^13^. Consistent predictors of ORV failure within the bacterial gut microbiota thus remain elusive.

Much of what we know about the mechanisms shaping oral vaccine performance comes from single-population studies focusing on individual risk factors. Here, we present a multicentre cohort study exploring the effect of maternal antibodies, EED biomarkers, and bacterial gut microbiota development on Rotarix response among infants in Malawi, India, and the UK.

## Results

We enrolled pregnant women during the third trimester in Blantyre (Malawi; n = 119), Vellore (India; n = 306), and Liverpool (UK; n = 60; **Table 1**). After delivery, infants received routine vaccines including two doses of Rotarix according to the national immunisation schedule (6 and 10 weeks in Malawi and India, 8 and 12 weeks in the UK). We measured rotavirus-specific IgA (RV-IgA) in maternal blood, cord blood, and breastmilk samples collected during or in the week after delivery, and in infant blood samples collected pre- and post-vaccination. In Indian participants, all serum samples were also assayed for rotavirus-specific IgG (RV-IgG). Six longitudinal stool samples were collected from each infant and assayed for rotavirus shedding, bacterial microbiota composition (16S rRNA V3–V4 sequencing), and/or EED biomarkers (**Figure 1A** and **Table S1**).

**Table 1.**
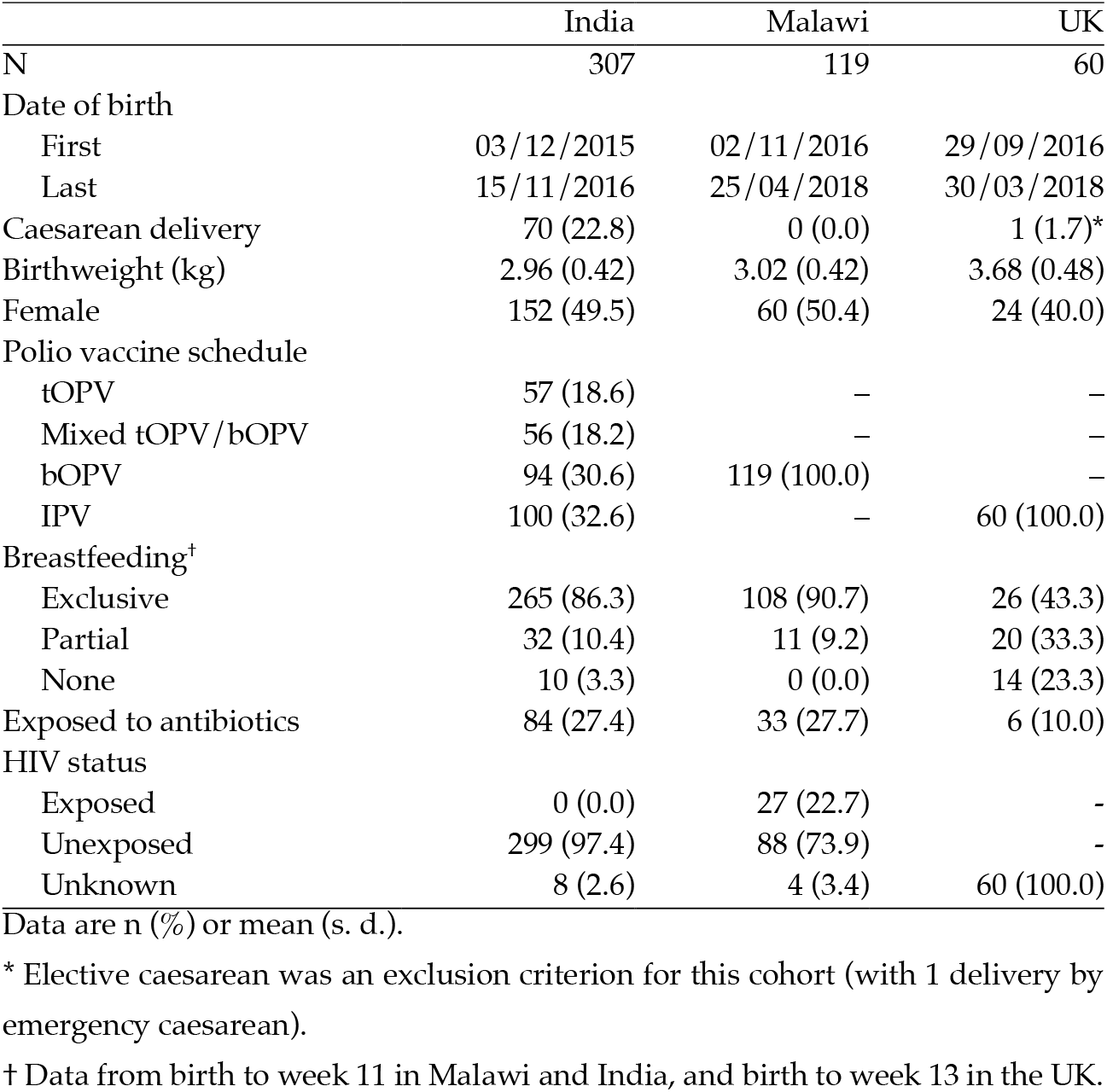
Baseline characteristics of study cohorts.

|  | India | Malawi | UK |
| --- | --- | --- | --- |
| N | 307 | 119 | 60 |
| Date of birth |  |  |  |
| First | 03/12/2015 | 02/11/2016 | 29/09/2016 |
| Last | 15/11/2016 | 25/04/2018 | 30/03/2018 |
| Caesarean delivery | 70 (22.8) | 0 (0.0) | 1 (1.7)* |
| Birthweight (kg) | 2.96 (0.42) | 3.02 (0.42) | 3.68 (0.48) |
| Female | 152 (49.5) | 60 (50.4) | 24 (40.0) |
| Polio vaccine schedule |  |  |  |
| tOPV | 57 (18.6) | – | – |
| Mixed tOPV/bOPV | 56 (18.2) | – | – |
| bOPV | 94 (30.6) | 119 (100.0) | – |
| IPV | 100 (32.6) | – | 60 (100.0) |
| Breastfeeding<sup>†</sup> |  |  |  |
| Exclusive | 265 (86.3) | 108 (90.7) | 26 (43.3) |
| Partial | 32 (10.4) | 11 (9.2) | 20 (33.3) |
| None | 10 (3.3) | 0 (0.0) | 14 (23.3) |
| Exposed to antibiotics | 84 (27.4) | 33 (27.7) | 6 (10.0) |
| HIV status |  |  |  |
| Exposed | 0 (0.0) | 27 (22.7) | – |
| Unexposed | 299 (97.4) | 88 (73.9) | – |
| Unknown | 8 (2.6) | 4 (3.4) | 60 (100.0) |

Data are n (%) or mean (s. d.).
\* Elective caesarean was an exclusion criterion for this cohort (with 1 delivery by emergency caesarean).
† Data from birth to week 11 in Malawi and India, and birth to week 13 in the UK.

**Figure 1.**
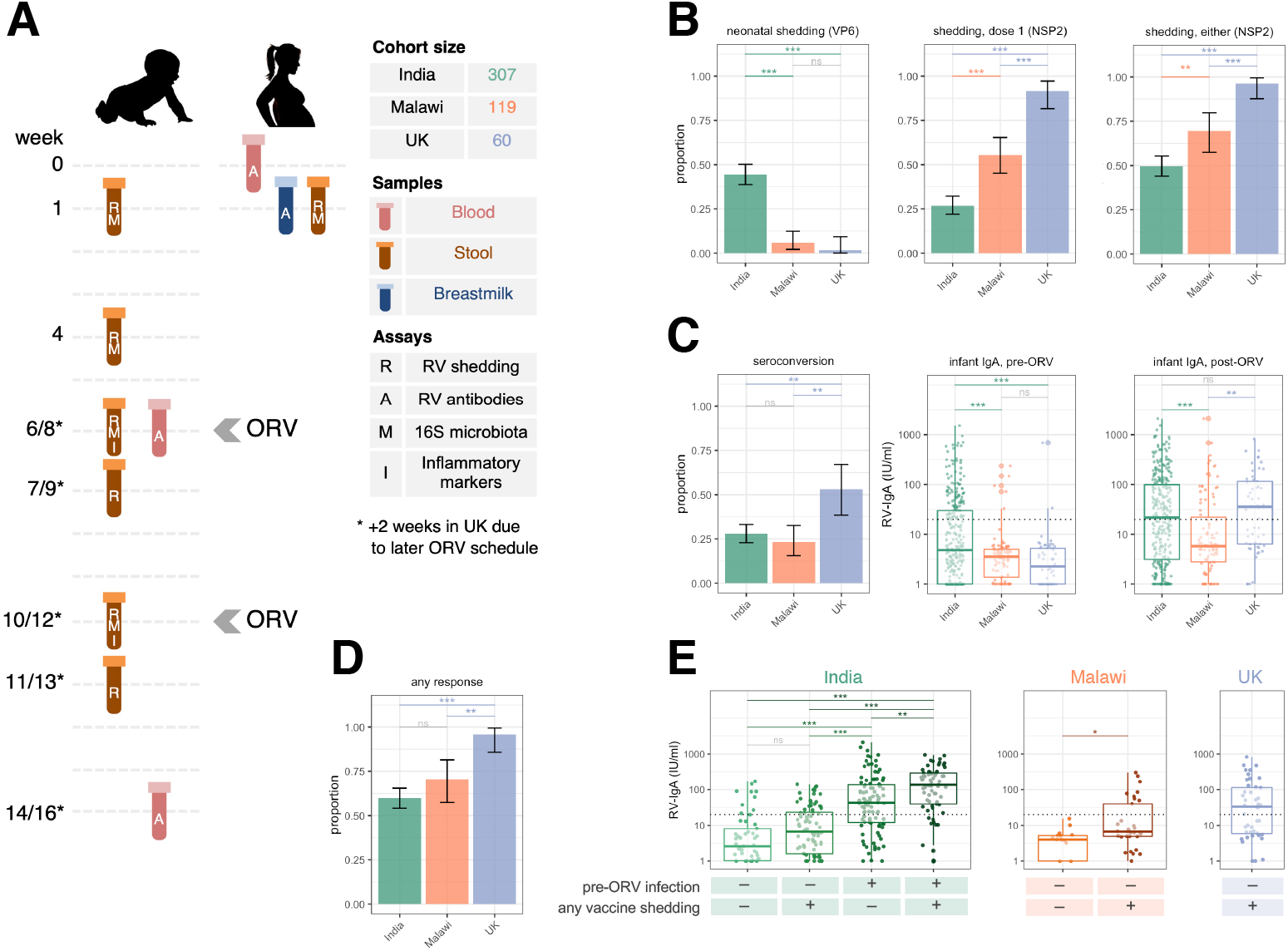
Study Design and Oral Rotavirus Vaccine Response. (A) Study design. (B to D) Geographic differences in (B) rotavirus shedding, (C) immunogenicity, and (D) cumulative vaccine response (seroconversion or shedding after either dose). Error bars represent Clopper–Pearson 95% confidence intervals. (E) Association between rotavirus shedding and post-vaccination antibody concentration. Gene targets for shedding assays are indicated in brackets. Groups were compared by Fisher’s exact tests with FDR correction (binary outcomes) or ANOVAs with post-hoc Tukey tests (continuous outcomes). The dotted lines at 20 IU/ml indicate the standard cut-off for RV-IgA seropositivity. ns, not significant; ORV, oral rotavirus vaccine; RV, rotavirus; * p <0.05; ** p <0.005; *** p <0.0005.

### ORV replication and immunogenicity

In the UK, we observed near-ubiquitous shedding of the first dose of ORV, with 55/60 (92%) shedding 1 week after vaccination and 24/54 (44%) continuing to shed after 1 month (**Figure S1A**). By contrast, dose 1 shedding was detected in 82/305 (27%) infants in India and 56/101 (55%) in Malawi (**Figure 1B**), and continued shedding after 1 month was much rarer in these cohorts (**Figure S1A**). ORV shedding after the second dose was also more common in the UK than India or Malawi, although discrepancies were less marked for this dose (**Figure 1B**). Shedding following at least one dose was observed in 54/56 (96%) infants in the UK, 151/304 (50%) in India, and 50/72 (69%) in Malawi (Fisher’s p values <0.005 for all between-country comparisons after false discovery rate [FDR] adjustment; **Figure 1B**).

Seroconversion was observed in 85/305 (28%) infants in India, 24/103 (23%) infants in Malawi, and 27/51 (53%) infants in the UK (**Figure 1C**). Geometric mean concentrations (GMCs) of RV-IgA (IU/ml) after vaccination were 20 (95% CI 16–25) in India, 9 (6–12) in Malawi, and 27 (17–45) in the UK (Tukey’s post-hoc p values <0.005 for comparisons between Malawi and other cohorts and 0.489 for India vs UK). Overall, at least one indicator of ORV response (seroconversion or shedding after at least one dose) was observed in 181/302 (60%) infants in India, 43/61 (70%) in Malawi, and 46/48 (96%) in the UK (**Figure 1D**).

### Neonatal rotavirus infection

A unique feature among Indian infants was the high rate of wild-type rotavirus shedding in the first week of life (136/306 [44%] based on qPCR, compared with 6/102 [6%] in Malawi and 1/58 [2%] in the UK; **Figure 1B**). We successfully characterised rotavirus genotype in 104 samples, of which 103 were positive for the strain G10P[11]. Overall, 166/304 (55%) infants in India had some indication of pre-vaccination rotavirus infection (week 1 shedding or baseline RV-IgA ≥20 IU/ml), whereas the corresponding rates were 10/90 (11%) in Malawi and 2/54 (4%) in the UK. Pre-vaccination infection was more common among infants born in tertiary care facilities in India (relative risk [RR] 1.98 [95% CI 1.72–2.19]; **Table S2)** and among infants delivered by caesarean section versus vaginal delivery (RR 1.31 [1.05–1.53]). All neonatal rotavirus infections were asymptomatic.

Consistent with the high prevalence of neonatal rotavirus infection, we observed significantly higher pre-vaccination RV-IgA concentrations in India compared with Malawi and the UK (**Figure 1C**). Prior infection was associated with a reduced likelihood of shedding ORV after at least one dose (RR 0.65 [0.47–0.83]; **Figure S1B** and **Table S3**). By contrast, pre-vaccination infection did not influence the likelihood of seroconversion (**Figure S1B** and **Table S4**), and where ORV replication was observed among previously infected infants, this significantly boosted post-vaccination RV-IgA concentrations (**Figure 1E**). Overall, our findings point to the high frequency and immunogenic nature of neonatal rotavirus infection in India. Indeed, despite the lower seroconversion rates in India compared with the UK, post-vaccination GMCs did not differ significantly between these countries (**Figure 1C**).

### Breastfeeding, growth, and sanitation

We examined several baseline health and demographic variables for their potential correlation with ORV response (**Tables S3** to **S5**). In India, seroconversion was positively correlated with exclusive breastfeeding (RR 2.04 [1.07–3.68]). Recruitment in this cohort spanned the global switch from trivalent OPV (tOPV) to bivalent OPV (bOPV) in April 2016. Seroconversion was more common in infants who received bOPV-only than tOPV-containing schedules (**Table S4**), although infants receiving tOPV were also more likely to be immunised during the dry season (January–May) given the timing of the tOPV–bOPV switch. Similar associations were apparent when considering post-vaccination RV-IgA concentration as an endpoint (**Table S5**). Moreover, RV-IgA was positively correlated with height-for-age Z score at the time of the first ORV dose, and was higher among infants living in houses built from permanent versus temporary or mixed materials (beta 2.03 [95% CI 1.26–3.26], where beta represents the estimated ratio of GMCs) or with access to treated water (beta 1.66 [1.04–2.67]). Baseline health and demographic variables were not strongly associated with ORV response in Malawi or the UK, although fewer covariates were measured in these cohorts.

### Maternal antibodies

Serum RV-IgA concentrations were significantly higher among mothers in Malawi than in India or the UK (**Figure 2A**; GMCs of 134 [120–150], 340 [256–452], and 186 [123–281] in India, Malawi, and the UK, respectively). Interestingly, maternal RV-IgA levels did not differ significantly between India and the UK, while breastmilk RV-IgA concentrations were significantly higher in Malawi and the UK than in India (GMCs of 25 [22–29], 97 [84–112], and 100 [68–146] in India, Malawi, and the UK, respectively). Thus, while the degree of maternal antibody exposure was greatest in Malawi, infants in the UK were by no means exempt from this potential ORV inhibitor.

**Figure 2.**
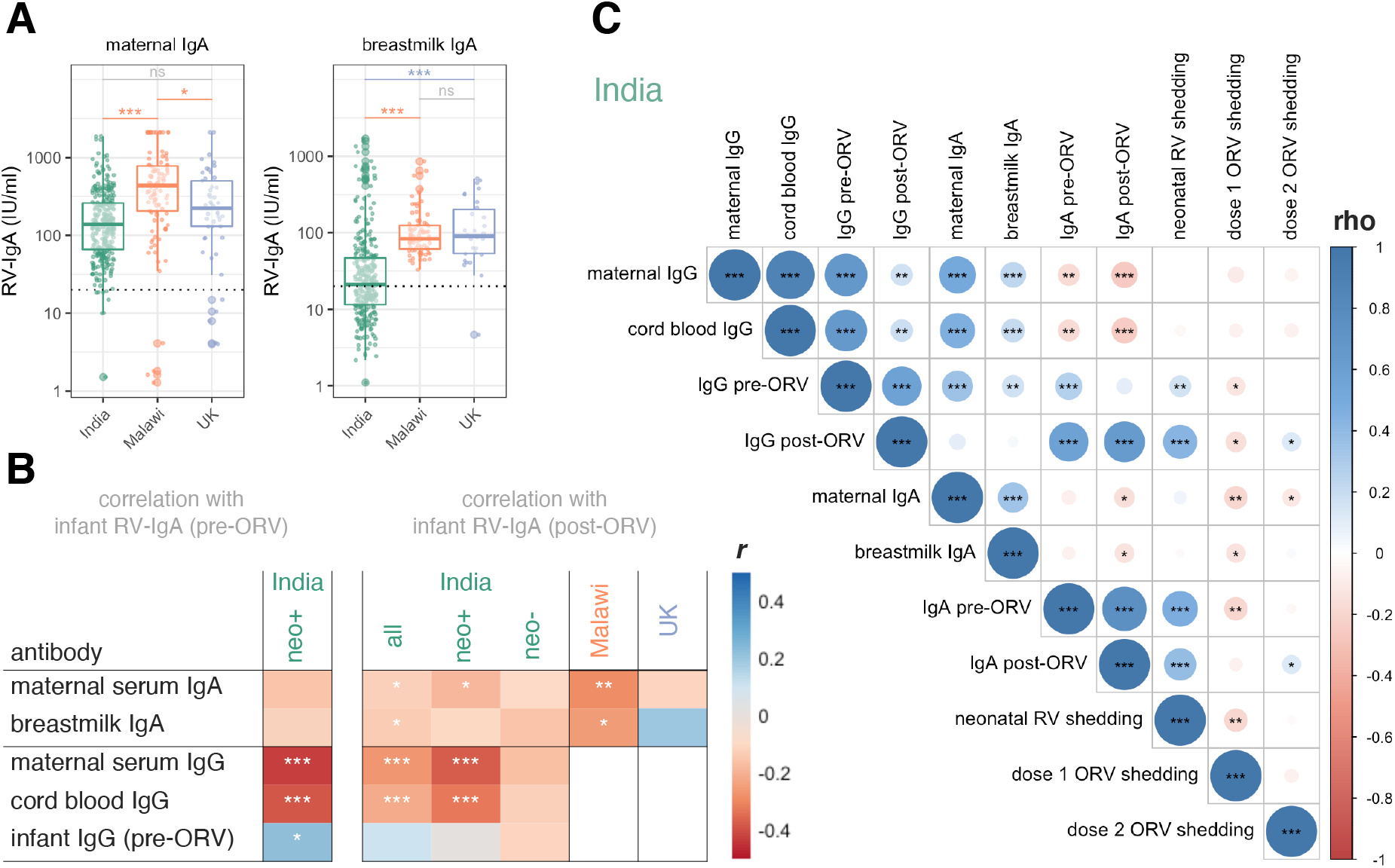
Association between Maternal Antibodies and Oral Rotavirus Vaccine Response. (A) Maternal antibody concentrations by cohort. Groups were compared by ANOVAs with post-hoc Tukey tests. The dotted lines at 20 IU/ml indicate the standard cut-off for RV-IgA seropositivity. (B) Association between maternal antibodies and RV-IgA formation. Log-transformed concentrations were compared using Pearson’s correlation coefficient (*r*). (C) Correlation between antibody concentrations and rotavirus shedding quantity (1/Ct) among Indian infants. Variables were compared using Spearman’s rank correlation coefficient (rho). neo+, infected with rotavirus pre-vaccination; neo-, uninfected with rotavirus pre-vaccination; ORV, oral rotavirus vaccine; RV, rotavirus; * p <0.05; ** p <0.005; *** p <0.0005.

Our analysis of the association between maternal antibodies and ORV outcome yielded several notable findings. First, maternal serum RV-IgA concentrations were negatively correlated with infant post-vaccination RV-IgA levels in each cohort (Pearson coefficient [*r*] of −0.29, −0.13, and −0.12 in Malawi, India, and the UK, respectively; **Figure 2B**). Although this correlation was not significant in the UK, this may reflect a lack of statistical power owing to the smaller size of this cohort, and we observed no significant evidence of effect modification between cohorts (p values of >0.05 for interaction terms between maternal RV-IgA and country). Serum IgA is not transmitted efficiently across the placenta and is therefore unlikely to directly influence ORV. However, we observed a strong correlation between maternal RV-IgA and cord blood RV-IgG in India (*r* 0.41, p <0.001), suggesting that maternal RV-IgA may be a valid (if imperfect) proxy for the degree of transplacental RV-IgG transfer (**Figure 2C**). Second, maternal serum RV-IgA levels were negatively correlated with ORV shedding in both India and Malawi (**Figure 2C** and **Table S3**). By contrast, ORV shedding in the UK was ubiquitous despite the fact that maternal antibody levels were equivalent to those in India. Finally, we observed no significant difference in maternal antibody levels according to week 1 rotavirus shedding status in Indian infants (**Table S2**), suggesting that neonatal G10P[11] viruses may not be subject to the same replication inhibition observed for ORV. However, among infants with pre-vaccination infection, we observed a strong negative correlation between maternal RV-IgG levels and infant RV-IgA at 6 weeks (*r* −0.42, p <0.001; **Figure 2B**).

Breastmilk RV-IgA was negatively correlated with ORV immunogenicity and/or shedding in India and Malawi (**Figure 2B** and **Tables S3** to **S5**). Disentangling the relative influence of breastmilk versus transplacental antibodies is challenging given the correlation between the two (**Figure 2C**). However, as noted above, ORV immunogenicity was higher in conjunction with exclusive breastfeeding in India – a finding at odds with a significant inhibitory effect of breastmilk antibodies on ORV response.

### Inflammatory biomarkers

We observed strong geographic discrepancies in EED biomarkers. Whereas α1-antitrypsin (α1AT; a marker of protein-losing enteropathy) was highest in Malawi and lowest in the UK, both myeloperoxidase (MPO; a marker of neutrophil activity) and α1 acid glycoprotein (a marker of systemic inflammation) were highest in Indian infants (**Figure S2A**). Despite their marked variation within and between cohorts, none of the biomarkers were significantly associated with seroconversion, ORV shedding, or post-vaccination RV-IgA (**Figure S2B** and **Tables S3** to **S5**).

### Geographic differences in microbiota development

We obtained high-quality faecal microbiota profiles in 2,086 samples (142,880 ± 136,113 [mean ± s.d.] reads per sample). Microbiota profiles were highly consistent across sequencing runs and facilities (explored by independently re-sequencing 10% of samples; **Figure S3**). Infant samples were dominated by a small number of taxa (**Figure S4**). The trajectory of microbiota development was highly distinct to each cohort, with significant deviations apparent as early as the first week of life. In particular, microbiota diversity and richness were significantly higher in Malawi than both other cohorts (**Figure 3A**), and numerous genera were enriched in Malawi compared with both India (21 genera with FDR p <0.1 in longitudinal mixed-effects models) and the UK (22 genera with FDR p <0.1; **Figure S5**). On the other hand, *Bifidobacterium, Enterococcus, Staphylococcus*, and *Streptococcus* were enriched in Indian infants compared with both other cohorts, while *Bacteroides, Citrobacter, Enterobacter, Klebsiella, Haemophilus, Lachnospiraceae*, and *Veillonella* were enriched in the UK (**Figures 3B** and **S5**). Several of these taxonomic discrepancies were also evident in maternal stool samples collected in the week after delivery (**Figure S4C**). Clustering of samples by country at the time of the first ORV dose was evident based on Bray–Curtis distances (**Figure 3C**). Moreover, using the machine-learning method Random Forests, samples could be accurately distinguished by country at all time-points (mean cross-validation accuracies of 83–98%; baseline accuracy 50%; **Figure 3D**). The genera most important to the predictive accuracy of these models differed between cohorts in both their prevalence and abundance (**Figures 3E** and **S6**). Indeed, simple cross-sectional comparisons of taxon prevalence were adept at selecting differential microbiota colonisation patterns between cohorts (**Figure S5C** and **Table S6**).

**Figure 3.**
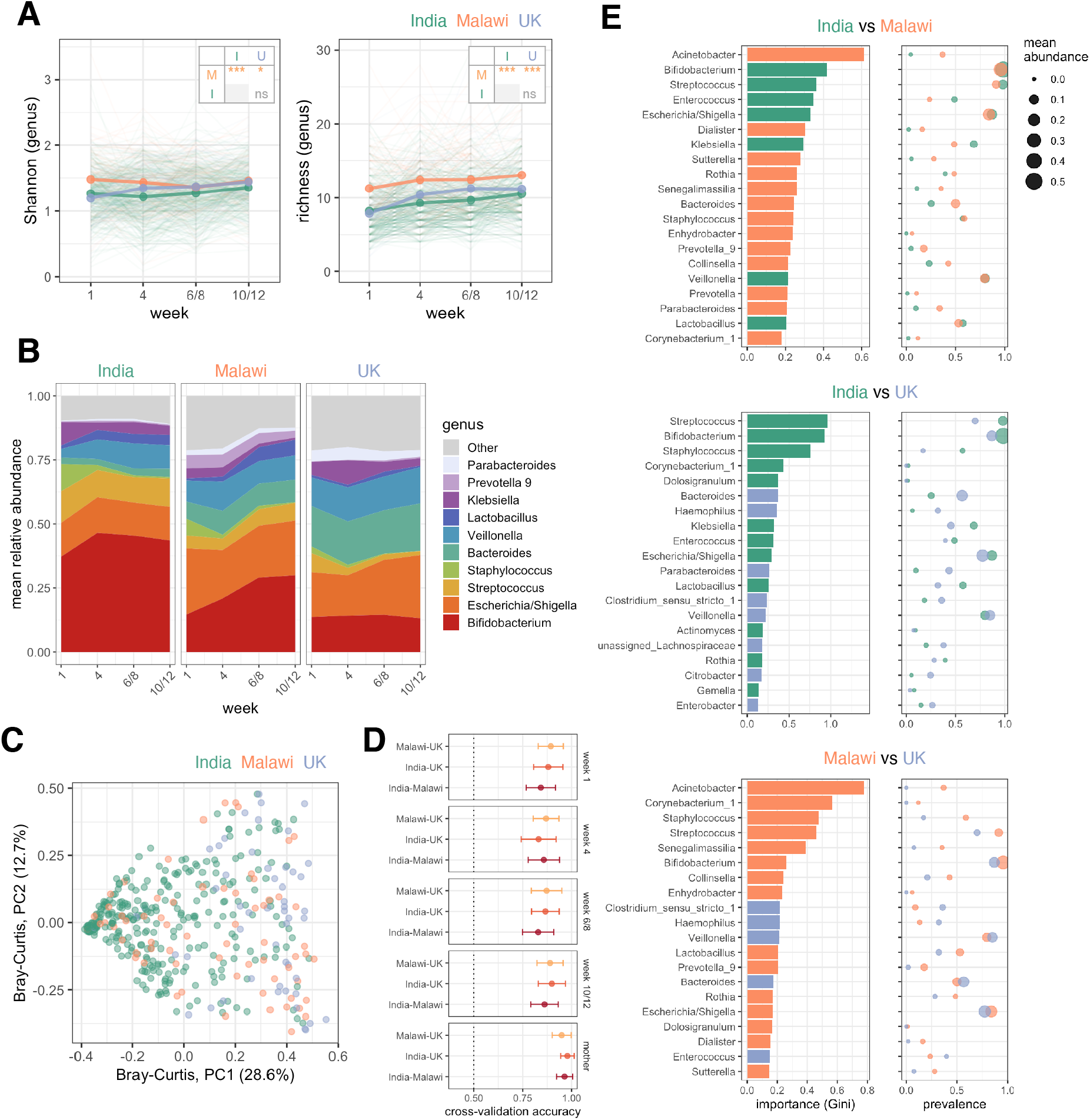
Geographic Differences in Microbiota Development. (A) Longitudinal analysis of alpha diversity. Points represent mean values. Groups were compared using longitudinal mixed-effects models including week as a covariate and study ID as a random effect. Pairwise comparisons between countries were FDR corrected. (B) Longitudinal plot of mean genus abundances. Genera were included if they were present with a mean relative abundance of ≥5% at one or more timepoints. (C) Principal coordinates analysis plot of weighted Bray-Curtis distances for samples collected before the first dose of ORV (week 6 in India and Malawi; week 8 in the UK). (D) Cross-validation accuracy of Random Forests models (mean ± s.d.). (E) The 20 most important genera selected by Random Forests for discriminating infants by country at the time of the first dose of ORV. Mean cross-validation importance scores based on Gini index are depicted alongside the prevalence (≥0.1% abundance threshold) and mean abundance of the corresponding genera in each country. PC, principal coordinate; ns, not significant; * p <0.05; *** p <0.0005.

### Microbiota composition versus ORV response

In both India and Malawi, we observed a significant negative correlation between microbiota diversity and ORV immunogenicity. In India, these discrepancies were apparent in longitudinal models (**Figure 4A**) and cross-sectional comparisons at the time of the first ORV dose (**Figure 4B** and **Table S4**). Interestingly, stratified analyses revealed the discrepancies to be specific to infants with no rotavirus exposure prior to receiving ORV (**Figure 4A**). A similar trend was evident in Malawian infants, albeit specifically at the time of the first dose of ORV (**Figure 4B** and **Table S4**). By contrast, microbiota diversity was not strongly associated with ORV immunogenicity in the UK. Neither Shannon index nor richness differed according to ORV shedding status in any cohort (**Figure S7** and **Table S3**).

**Figure 4.**
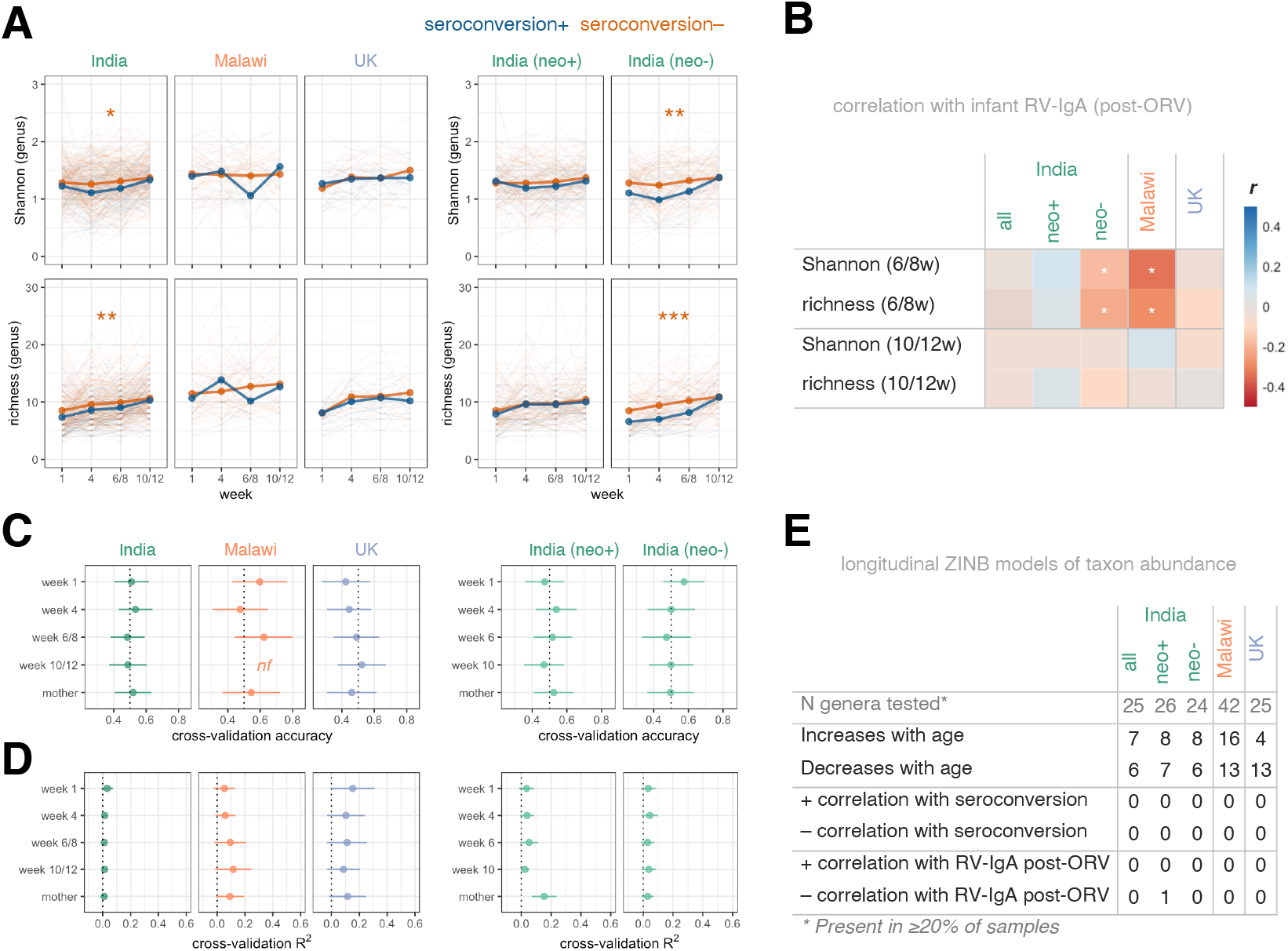
Association between Microbiota Development and Oral Rotavirus Vaccine Response. (A) Longitudinal analysis of microbiota diversity. Points represent mean values. Groups were compared using longitudinal mixed-effects models including week as a covariate and study ID as a random effect. (B) Association between microbiota diversity and RV-IgA formation. Variables were compared using Pearson’s correlation coefficient (*r*). Microbiota richness values (number of genera detectable at ≥0.1% abundance) were log-transformed. (C and D) Random Forests cross-validation accuracy for (C) seroconversion and (D) RV-IgA concentrations based on genus abundance data (mean ± s.d.). (E) Longitudinal zero-inflated negative binomial models of genus abundance. Genera with FDR-adjusted p values of <0.1 are highlighted. neo+, infected with rotavirus pre-vaccination; neo-, uninfected with rotavirus pre-vaccination; nf, not fitted as <10 responders; ORV, oral rotavirus vaccine; RV, rotavirus; ZINB, zero-inflated negative binomial; * p <0.05; ** p <0.005; *** p <0.0005.

In spite of the significant differences in microbiota diversity, we did not observe clear differences in taxonomic composition according to ORV response. Cross-sectional Random Forests models based on genus abundances failed to accurately predict immunogenicity (**Figures 4C** and **4D**) or shedding status (**Figures S7C** and **S7D**). While numerous genera either increased or decreased in abundance with age, very few exhibited differential abundance according to ORV outcome when assessed via longitudinal models (**Figures 4E** and **S7E**). Exceptions include *Sutterella*, which was negatively correlated with dose 1 ORV shedding in Malawian infants, while *Lactobacillus, Corynebacterium*, and *Dolosigranulum* were negatively correlated with ORV shedding and/or final RV-IgA concentration in India (**Table S7**). Overall, while microbial exposure and colonisation appears to be significantly higher in infants who fail to respond to ORV, we did not observe consistent discrepancies in specific bacterial taxa according to vaccine outcome.

### Multivariate analysis

To delineate the contribution of different risk factors in shaping ORV response, we performed an integrated analysis based on three modules of input data: (i) demographic and baseline health variables (n = 18; see **Table S4** for complete list); (ii) exposure/antibody data (n = 12), including EED biomarkers and maternal antibody concentrations; and (iii) pre-vaccination microbiota composition, comprising genus abundances at the time of the first ORV dose (n = 126). We used the machine-learning algorithm Random Forests to predict ORV outcome in Indian infants (the largest cohort in terms of number of infants and number of recorded variables) using each module in turn and all modules combined. The prediction of seroconversion and ORV shedding was poor for all modules (**Figure 5A**). Post-vaccination RV-IgA concentration was predicted with modest accuracy by exposure/antibody data (cross-validation R^2^ of 0.36 ± 0.11 [mean ± s.d.]), and this module alone offered predictive accuracy commensurate with the integrated analysis combining all variables (R^2^ of 0.38 ± 0.10; **Figure 5A**). Consistent with the univariate analyses described above, the most important variables underlying the prediction of post-vaccination RV-IgA levels were pre-vaccination rotavirus infection and maternal RV-IgG concentration (**Figure 5B**).

**Figure 5.**
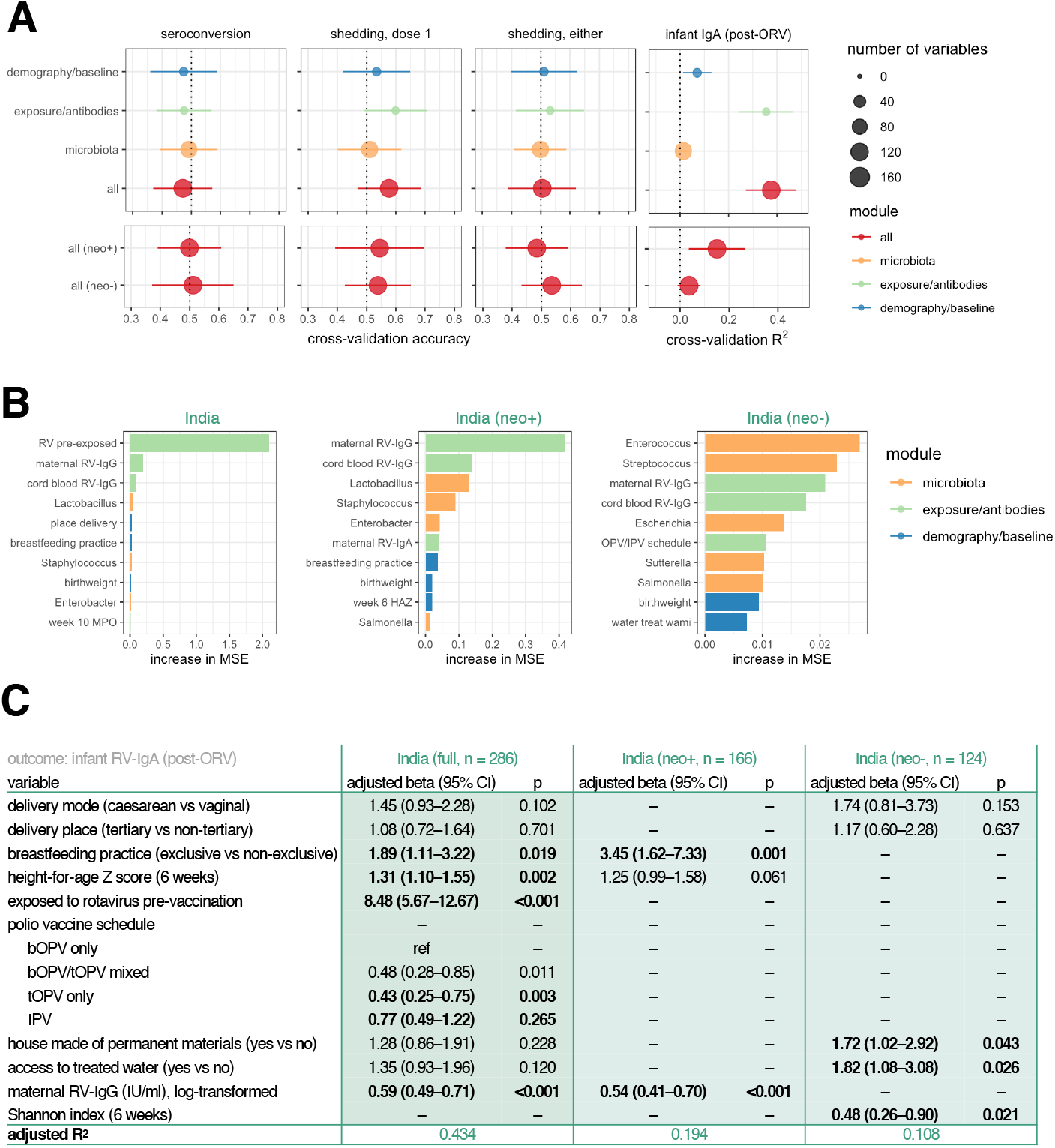
Integrated Analysis for Prediction of Oral Rotavirus Vaccine Response. (A) Random Forests models cross-validation accuracy for prediction of vaccine outcome based on demographic, exposure/antibody data, and pre-vaccination (week 6) microbiota data (mean ± s.d.). (B) The top 10 variables selected by Random Forests for prediction of RV-IgA concentration based on mean cross-validation importance score. (C) Multivariate regression models for post-vaccination RV-IgA. Variables were included if significantly associated with RV-IgA concentration (p <0.05) during univariate analyses. See **Table S5** for full univariate and multivariate outputs. neo+, infected with rotavirus pre-vaccination; neo-, uninfected with rotavirus pre-vaccination; MSE, mean squared error; ORV, oral rotavirus vaccine; RV, rotavirus.

We complemented the machine-learning models with multivariate regressions. Covariates with a p value of <0.05 in univariate analyses were selected for inclusion in these models (**Tables S3** to **S5**). Notably, this approach revealed breastfeeding practice, linear growth, pre-vaccination exposure, and polio vaccine schedule to be significantly correlated with post-vaccination RV-IgA (**Figure 5C**). Associations varied according to pre-vaccination exposure status – whereas breastfeeding practice and maternal antibody levels were strong predictors of RV-IgA among infants with neonatal rotavirus exposure, microbiota diversity, house type, and access to treated water were significantly associated with vaccine immunogenicity in those without prior exposure. Overall, the multivariate regressions offered a predictive accuracy commensurate with or exceeding that of Random Forests (R^2^ of 0.43 in the complete cohort and 0.11–0.19 in the stratified analyses for RV-IgA regressions).

## Discussion

ORV is unique among oral vaccines in being routinely administered in both high-income countries and LMICs. As such, the vaccine provides a valuable tool for probing the divergent trajectories of the developing immune system in different settings. Here, we report on the extent to which several key determinants of infant immune development differ among three disparate populations, and the degree to which these factors shape ORV response.

As expected, Rotarix response was significantly impaired among infants in Malawi and India. While the later vaccination schedule in the UK (8/12 weeks vs 6/10 weeks) may contribute to these differences by providing more time for passively acquired maternal antibodies to wane, overall shedding rates after both doses in Malawi and India still fell short of the near-ubiquitous dose 1 shedding observed in the UK, highlighting the barrier to ORV replication that emerges within the first months of life in LMICs. Seroconversion rates in India and Malawi were broadly consistent with those previously reported in LMICs^14,15^, but lower than expected in the UK^16^. These findings highlight the suboptimal nature of RV-IgA as a correlate of protection. While higher RV-IgA concentrations correlate with a lower risk of subsequent rotavirus infection, these associations often fail to hold when considering protection against rotavirus-associated gastroenteritis^17^. We advocate the further use of vaccine virus replication as an adjunctive measure of ORV response in future trials.

For a risk factor to account for broad geographic trends in ORV efficacy, it is reasonable to expect it to be more common or observed at higher levels in LMICs than high-income countries. This was not the case for several of the factors examined in this study. Maternal serum RV-IgA levels were comparable in the UK and India (although significantly higher in Malawi), while breastmilk RV-IgA levels were higher in the UK than in India. These results are somewhat surprising given that recent rotavirus exposure and consequent boosting of antibody levels might be expected to occur more frequently among mothers in LMICs with a high rotavirus disease burden. Moreover, in previous studies, RV-IgA concentrations were significantly higher in India than the USA, with intermediate levels in South Africa^18,19^. Our findings caution against the broader extrapolation of these trends. The high maternal RV-IgA levels in the UK may reflect the immunogenic nature of early rotavirus exposures and the subsequent boosting of antibodies by mild or asymptomatic re-infection ^20^.

Nonetheless, our findings confirm the potential inhibitory effect of maternal antibodies on ORV among infants in LMICs. Whereas previous studies of this phenomenon have typically focused on immunogenicity endpoints^8,21^, we observed a reduction in both shedding and immunogenicity, suggesting that the effect of maternal antibodies is mediated in part through a reduction in ORV replication efficiency. This could potentially involve the transudation of transplacental RV-IgG antibodies across the mucosal epithelium or the direct neutralisation of vaccine viruses by RV-IgA in breastmilk. In neonatal macaques, passively acquired monoclonal IgG antibodies have been shown to confer protection against lentivirus infection at mucosal surfaces^22^, and a similar mechanism may influence ORV. The presence of a leaky mucosal barrier following early-life enteropathogen exposure could feasibly enhance the transfer of maternal RV-IgG to the gut lumen among infants in LMICs^23^ and thereby enhance their inhibitory effect, although this possibility remains untested. If breastmilk RV-IgA is involved in the inhibition of ORV replication, the effect does not appear to be mitigated by the temporary withholding of breastfeeding, which failed to boost ORV response in controlled trials^24,25^. Our findings are also consistent with the inhibition of downstream antigen processing by transplacental antibodies, as evidenced by the strong inverse correlation between maternal RV-IgG and infant RV-IgA formation in Indian infants with neonatal rotavirus infection. Overall, one could reasonably envision a combination of upstream inhibition of ORV replication (via RV-IgA and/or transudated RV-IgG) and downstream inhibition of antigen processing (via circulating RV-IgG) that synergistically inhibit ORV outcome. However, these mechanisms alone are clearly insufficient to prevent a robust ORV response given that the vaccine was ubiquitously shed and was highly immunogenic in the UK, where maternal antibody levels were high and >75% of infants were breastfed.

We considered several biomarkers as a proxy for EED. Although α1AT concentrations were significantly lower in the UK than both other cohorts, MPO levels were similar in the UK and Malawi, calling into question the extent to which this marker accurately captures the onset of EED. We did not observe a significant relationship between any of the measured EED biomarkers and ORV outcome, in keeping with the growing literature in this field^11,26^. Together, these findings imply either that EED is not a primary driver of oral vaccine failure or that the condition is not accurately captured by the current suite of faecal and systemic biomarkers (quite possibly both).

We observed marked geographic discrepancies in composition of the bacterial microbiota. Microbiota diversity was significantly higher among Malawian infants, while Indian infants could be distinguished by their high *Bifidobacterium* abundance. The latter is interesting given that *Bifidobacterium* has previously been suggested to enhance immunological memory responses to a range of vaccine targets in Bangladesh^27^, yet this genus was least abundant among infants in the UK that were most responsive to ORV. Similarly, we failed to recapitulate other associations that have between documented between bacterial microbiota composition and ORV outcome, such as the higher abundance of Bacteroidetes among non-responders in Ghana^12^. However, we observed a significant negative correlation between microbiota diversity and ORV immunogenicity – apparent among Indian infants throughout infancy and in Malawian infants specifically prior to the first vaccine dose. Although this is the first study to find such an association for ORV, we previously reported a negative correlation between microbiota diversity and monovalent type 3 OPV response among 6–11 month-old Indian infants^28^. Several indirect explanations for this trend are plausible. For example, microbiota richness may act as a proxy for other early-life exposures that shape ORV outcome, such as non-polio enteroviruses^29^. The fact that ORV immunogenicity was impaired among Indian infants in households without access to treated water would support this notion. It is also possible that exposure to greater microbial diversity may foster a state of hyporesponsiveness at the mucosal epithelium that impairs oral vaccine outcome. When viewed in this way, impaired vaccine response could be seen as a counterpart to hyperresponsive immune states associated with low early-life microbiota diversity, such as atopic disease^30^. Notably, low microbiota diversity is often used as a marker of ‘dysbiosis’^31^, albeit largely based on studies in high-income settings. A clear understanding of the signatures of ‘healthy’ microbiota development in different settings is lacking. Our findings allude to a more complex narrative whereby such signatures vary not only by geographic setting and age, but by the health outcome being considered.

Our study has several limitations. As noted above, RV-IgA is a suboptimal correlate of protection against rotavirus-associated gastroenteritis. We compensated for this by considering multiple endpoints, including seroconversion and post-vaccination shedding. Whilst the potential contribution of batch effects to the observed differences between populations cannot be completely ruled out, we went to considerable lengths to ensure reproducibility and comparability across sites, using the same reagent lots and standards. In the case of microbiota composition, we independently re-sequenced 10% of samples at a separate facility and recapitulated the original microbiota composition with high accuracy. Finally, profiling the virome, proteome, transcriptome, and metabolome can now be achieved using small sample volumes and would provide valuable insight into systemic immune status at the time of vaccination in future studies^32^.

Notwithstanding these caveats, this study advances our understanding of the potential mechanisms influencing ORV response in several ways. By considering how several risk factors of ORV failure differ within and between populations, we have been able to place these mechanisms within a broader global context. Our study confirms the inhibitory effect of maternal antibodies on ORV immunogenicity in LMICs, and suggests this is mediated partly by a reduction in ORV replication. We observed that asymptomatic neonatal rotavirus exposure was strongly associated with RV-IgA formation, providing further support for the potential of neonatal vaccination as a pragmatic approach to achieve greater rotavirus vaccine impact. Finally, while specific bacterial taxa do not appear to drive within-population differences in ORV response, high microbial exposure in early life may contribute to the impaired efficacy of this vaccine in LMICs.

## Data Availability

The raw sequence data generated during this study have been deposited in the European Nucleotide Archive (accession number PRJEB38948). Processed data and analysis code generated during this study are available on github (https://github.com/eparker12/RoVI).

## Acknowledgements

We thank all members of the clinical study teams in Vellore, Blantyre, and Liverpool, including Falak Diab, Siobhan Holt, and the research midwives at the Liverpool Women’s Hospital; Dawn Redman and the team of research nurses at Alder Hey Children’s Hospital; Uma Raman, Charlet, Margaret, Jacklin, and the field research assistants at Christian Medical College, Vellore; and James Tamani, Anna Ainani, Amisa Chisale, Bertha Masamba, Carlo Gondwe, and Evelyn Gondwe in Blantyre, Malawi. The UK and Malawi sites were funded by the UK Medical Research Council and the UK Department for International Development (Newton Fund MR/N006259/1). M.I.G. is partly supported by the NIHR HPRU in Gastrointestinal Infections. K.C.J. is funded by a Wellcome International Training Fellowship (number 201945/Z/16/Z). The site in India was funded by the Government of India’s Department of Biotechnology. Richard Eccles, Anita Lucaci, Richard Gregory, John Kenny, and other staff at the Centre for Genomic Research (University of Liverpool) provided valuable support for the 16S microbiota sequencing work, as did Laurence Game at the Imperial BRC Genomics Laboratory. Above all, we are grateful to the families involved in the study.

## Author contributions

Conceptualisation, M.I.G., N.A.C., A.C.D., N.C.G., and G.K.; Methodology, C.B., E.P.K.P., A.C.D., M.I.G., I.P., and S.B.; Software, E.P.K.P.; Validation, E.P.K.P.; Formal Analysis, E.P.K.P.; Investigation, C.B., E.P.K.P., N.C.-V., D.H., J.L., S.B., I.P., S.G., S.I., V.K.S., B.B., S.S., B.S.R., J.M., and E.C.; Resources Provision, K.N.S., A.G., V.P.V., Q.D., N.C., M.T., and N.A.C.; Data Curation, C.B., E.P.K.P., K.N.S., S.V., J.M., and M.I.G.; Writing – Original Draft, E.P.K.P., C.B., A.C.D., S.B., I.P., A.D., M.I.G., and N.C.G.; Writing – Review & Editing, B.K., Visualisation, E.P.K.P; Supervision, N.A.C., A.C.D., K.C.J., N.C.G., B.K.; Project Administration, C.B., M.I-G., I.P., S.B., K.N.S., K.C.J.; Funding Acquisition, M.I.G., G.K.

## Declaration of Interests

M.I.G. has received research grants from GSK and Merck, and has provided expert advice to GSK. K.C.J. and N.A.C. have received investigator-initiated research grant support from GSK. N.A.C. has received research grant support and honoraria for participation in rotavirus vaccine data safety monitoring committee meetings from GSK.

## Supplemental figure titles and legends

**Figure S1. Association between pre-vaccination rotavirus exposure and oral rotavirus vaccine response**. (A) Rotavirus shedding across all timepoints based on VP6-specific PCR. Groups were compared by Fisher’s exact tests with FDR correction. (B) Association between pre-vaccination rotavirus exposure and ORV outcome in Indian infants. Prior exposure was defined by week 1 rotavirus shedding or detection of RV-IgA at baseline (≥20 IU/ml). Exposed and unexposed groups were compared by ANOVA (log antibody concentrations) or Fisher’s exact tests (shedding/seroconversion). The cohorts from Malawi and the UK are displayed for reference; statistical comparisons among cohorts are shown in **Figure 1**. Error bars represent Clopper–Pearson 95% confidence intervals. ns, not significant; ORV, oral rotavirus vaccine; RV, rotavirus; *, p <0.05; **, p<0.005; ***, p <0.0005.

**Figure S2. Association between environmental enteric dysfunction and oral rotavirus vaccine response**. (A) Environmental enteric dysfunction biomarkers by cohort. Groups were compared by ANOVAs with post-hoc Tukey tests. (B) Association between inflammatory biomarkers and RV-IgA formation. Log-transformed concentrations were compared using Pearson’s correlation coefficient (*r*). α1AT, α1-antitrypsin; α1AG, α1 acid glycoprotein; MPO, myeloperoxidase; neo+, infected with rotavirus pre-vaccination; neo-, uninfected with rotavirus pre-vaccination; ORV, oral rotavirus vaccine; RV, rotavirus.

**Figure S3. Replicability of microbiota sequencing across runs and facilities**. Positive controls including an infant stool (BSctrl), maternal stool (MSctrl), and a mock bacterial community (MCctrl) were included in every plate, with up to up to four plates per run. Alpha diversity (A) and beta diversity (B) for these positive controls were highly replicable from run to run, with sample type accounting for the majority of variation among controls based on linear regression (for alpha diversity) and PERMANOVA (for beta diversity). A minimum detection threshold of 0.1% abundance was used for richness and unweighted Bray–Curtis metrics. (C) Proportion of variation associated with run for each sample group. R^2^ and statistical significance were determined separately for each sample group by PERMANOVA. Mean R^2^ values are indicated by dotted lines. (D–F) For 10% of samples (90 week-1 infant samples, 90 week-10/12 infant samples, and 90 maternal samples evenly distributed across the study sites), we validated the microbiota pipeline at a separate sequencing facility in London. PCR, quantification, and pooling all involved the same methods and reagents (ordered separately), although sequencing was performed via Illumina MiSeq rather than HiSeq (used for the majority of runs in Liverpool). The order of samples during the 10% validation was randomised across three plates. Microbiota composition was highly replicable based on alpha diversity (D), beta diversity (E), and relative abundance of major genera (F), with sample ID accounting for >97% of variation based on linear regression or PERMANOVA. In (B), replicates are linked by a line, although often the overlap is so precise that these lines are indistinguishable. The proportion of variation associated with sample code was equivalent when using unweighted distances (R^2^ = 0.996). PC, principal coordinate; * p<0.05.

**Figure S4. Genus profiles of infant and maternal samples**. (A) Hierarchical clustering of stool samples (columns) based on presence/absence of common genera (rows). Genera detected in 1% of samples were included. Rows and columns were clustered using Ward’s minimum variance hierarchical clustering method. (B) Genus abundance versus prevalence for infant samples. Samples in each cohort were generally dominated by a small number of taxa with high prevalence and abundance. All infant samples from each cohort were included in the prevalence and abundance calculations (week 1, week 4, week 6/8, and week 10/12). Margins display density plots. (C) Geographic discrepancies in maternal microbiota composition, including comparisons of alpha diversity (left panel) and genus composition (right panel). Diversity metrics were compared using ANOVAs with post-hoc Tukey tests. Only genera from **Figure 4B** are shown. ns, not significant; **, p <0.05; **, p <0.005; ***, p <0.0005.

**Figure S5. Discriminant genera by country**. (A) Longitudinal relative abundance plots for major genera (mean abundance ≥5% in at least one country for one or more timepoints). Lines show local weighted regression (loess) fits with 95% confidence intervals. (B) Mixed-effects zero-inflated negative binomial models of genus abundance. Genera were included if present in at least 20% of samples from at least one country being compared. Study ID was included as a random effect. Genera with an FDR-adjusted p values of <0.1 are highlighted. (C) Cross-sectional comparisons of genus prevalence. A minimum detection threshold of 0.1% abundance was used to define the presence/absence of a taxon. Genera with an FDR-adjusted p values of <0.1 are highlighted.

**Figure S6. Top 20 genera for prediction of country using Random Forests**. See **Figures 4E** and **4F** for additional details.

**Figure S7. Association between microbiota development and oral rotavirus vaccine response**. (A and B) Longitudinal analysis of microbiota diversity in relation to (A) dose 1 shedding status and (B) shedding after either dose. Points represent mean values. Groups were compared using longitudinal mixed-effects models including week as a covariate and study ID as a random effect. (C and D) Random Forests cross-validation accuracy for (C) dose 1 shedding and (D) shedding after either dose based on genus abundance data (mean ± s.d.). (E) Longitudinal zero-inflated negative binomial models of genus abundance. Genera with FDR-adjusted p values of <0.1 are highlighted, with further details provided in **Table S7**. (D) Number of samples per cross-validation fold during cross-sectional Random Forests models. All available samples were included in models for post-vaccination RV-IgA concentration. For binary outcomes, we randomly selected 50 infants per group in each iteration of cross-validation, or the number of samples in the minority group if this was <50. neo+, infected with rotavirus pre-vaccination; neo-, uninfected with rotavirus pre-vaccination; nf, not fitted as <10 samples in minority group; ORV, oral rotavirus vaccine; RV, rotavirus; ZINB, zero-inflated negative binomial.

**Figure S8. Removal of extraction batches with potential cross-contamination**. (A) Read counts in no-template and extraction controls. All no-template controls were clear of substantial amplification (left panel), as were pooled extraction controls from Malawi and the UK (middle panel). Four pools of extraction controls from India yielded significant amplification (>10,000 sequences). When the individual extraction controls from these pools were sequenced (right panel), two gave significant amplification (corresponding to extraction batches 14 and 27). (B) Beta diversity plots and (C) genus composition of these extraction controls revealed two clusters consistent with contamination of extraction batches 14 and 27. We therefore removed the samples associated with these extraction batches from the final analysis (n = 8). NTC, no-template PCR control; PC, principal coordinate.

## Methods

### Study design

Pregnant women were enrolled during the third trimester at three study sites: Blantyre (Malawi), Vellore (India), and Liverpool (UK). Women were included if they provided informed written consent to participate in the study and were willing to stay in the study area for 4 months following delivery. Exclusion criteria included: congenital immune deficiency; chronic renal or liver failure; other chronic illnesses which may affect immune function; non-singleton pregnancy; low birthweight or pre-term birth (<34 weeks gestation); congenital anomalies and other neonatal complications requiring prolonged hospitalisation; and delivery by elective caesarean (UK only). Mothers provided a venous blood sample as well as a cord blood sample during birth and a stool sample during the week after delivery. Infants provided two blood samples and four stool samples over the course of the study (**Figure 1A**). The study was approved by the Institutional Review Board at the Christian Medical College (CMC) in Vellore, the College of Medicine Research and Ethics Committee in Blantyre, and the North West – Liverpool East Research Ethics Committee in Liverpool. The trial is registered with the Clinical Trials Registry of India (CTRI/2015/11/006354) and the study protocol has previously been published^33^.

Routine vaccines were administered according to the routine schedule at each site, including OPV at 0, 6, and 10 weeks of age in Malawi and India. In Malawi, all infants were born after April 2016 – the date of the global switch from tOPV to bOPV. In India, 98/307 (32%) infants were born before April 2016. Infants therefore received tOPV (57/307 [19%]), a mixed schedule of tOPV and bOPV (56/307 [18%]), or bOPV (94/307 [31%]). To explore the potential inhibitory effect of OPV on ORV^34^, the 6 and 10 week doses of OPV were replaced with inactivated poliovirus vaccine (IPV) in a sequentially recruited cohort in India (100/307 [33%]). The comparison of OPV and IPV arms is described elsewhere (Babji et al, in preparation). Briefly, while shedding after the first dose of ORV was less common in OPV than IPV recipients, cumulative shedding after both doses was comparable in the two arms, as were seroconversion rates and post-vaccination RV-IgA levels. Given the lack of association between study arm and ORV immunogenicity, we pooled the IPV and OPV arms in the present study.

### Sample processing and storage

Whole blood was collected in anti-coagulation EDTA-tubes (BD) and stored at 4°C for up to 12 hours until collection by laboratory staff. Fractions were separated by centrifugation at 1,500–2,000 g in a benchtop centrifuge (Eppendorf). A disposable plastic transfer pipette was used to aspirate the plasma down to ∼1 mm from the red blood cells. The plasma was aliquoted into two screw cap cryo-tubes (Starlab). The buffy coat and red blood cells were subsequently collected into separate cryovials and all samples were stored at −70°C.

Stool and breastmilk samples were collected in sterile sample pots by participants and were shipped to the respective laboratory by courier within 24 hours in the UK and within 4 hours in India and Malawi (to account for ambient temperatures). Samples were kept at 4°C for a maximum of 8 hours until processing. Breastmilk samples were stored in 2 ml aliquots in SuperLock tubes (Starlab) at −70^°^C for maternal RV-IgA analysis. Two 10% stool suspensions were prepared in sterile phosphate-buffered saline (PBS) for assessing rotavirus shedding. Further aliquots of neat stool were stored in 2 ml SuperLock tubes (Starlab) at −70°C for microbiota analysis and inflammatory biomarker measurement. Stool samples were stored at −70°C for a maximum of 2 weeks prior to DNA extraction for microbiota analysis.

### Rotavirus-specific antibodies

RV-IgA and RV-IgG laboratory assays were conducted at CMC, Vellore, for samples from all three study sites using a custom antibody-sandwich ELISA ^35^. Briefly, 96-well plates (Costar) coated with rabbit hyperimmune serum to rotavirus were incubated with purified cell culture lysates (WC3) or mock-infected MA104 cells. Serial dilutions of standard and test sera were added followed by biotinylated rabbit anti-human IgA (Jackson ImmunoResearch Laboratories) for detection of RV-IgA and biotinylated rabbit anti-human IgG (Vector Laboratories). Absorbance was subsequently read at 492 nm. Background-corrected optical density values from sample wells were compared with the standard curve and IgA or IgG concentration was determined based on derived units of IgA or IgG arbitrarily assigned to the respective standard curve, with a minimum detection limit of 1 IU/ml. Seroconversion was defined as detection of RV-IgA at ≥20 IU/ml in previously seronegative infants or a 4-fold increase in RV-IgA concentration among infants who were seropositive at baseline.

### Rotavirus shedding

Quantification of rotavirus shedding was performed at each study site. RNA was extracted using the RNeasy mini kit (Qiagen) following the manufacturer’s instructions. Briefly, 10% PBS stool suspensions were defrosted on ice, vortexed, and briefly centrifuged (10,000 g for 10 seconds) to pellet larger debris. 250 μl of β-ME in buffer RTL was added to 250 μl of supernatant. The mixture was then vortexed before the addition of 250 μl of 70% molecular-grade ethanol. The RNeasy Mini Handbook (5^th^ Edition, October 2013) was followed for subsequent steps, and RNA was eluted in 50 μl RNase-free water. cDNA was generated immediately after RNA extraction. The master mix for cDNA conversion comprised: 7 μl of 10x Taq DNA PCR buffer (Invitrogen), 7 μl of 50 mM MgCl_2_ (BIOLINE), 1 μl of Random Primers (Invitrogen), 2 μl of 10 mM dNTP (Invitrogen), 1 μl of DTT (Invitrogen), 2 μl of SuperScript III reverse transcriptase (Invitrogen), and 10 μl of RNase-free water (Invitrogen). 40 μl of eluted RNA was denatured at 95°C for 5 minutes and subsequently cooled on ice for 2 minutes. 30 μl of master mix was added to each RNA sample, mixed gently by pipetting, and centrifuged briefly. The tube was placed in a thermal cycler and incubated at 25°C for 10 minutes, 37°C for 1 hour, then 95°C for 5 minutes. The cDNA was stored at −20°C.

Previously validated primers and probes were used for the qPCR-based detection of rotavirus NSP2 ^36^ and VP6 ^37^ in two separate reactions. cDNA and reagents were thawed on ice. The master mix for qPCR comprised: 12.5 μl of Platinum qPCR supermix-UDG 2x (Invitrogen), 2 μl of 10 μM forward primer (5’-GAACTTCCTTGAA-TATAAGATCACACTGA-3’ [NSP2] or 5’-GACGGVGCRACTACATGGT-3’ [VP6]), 2 μl of 10 μM reverse primer (5’-TTGAAGACGTAAATGCATACCAATTC-3’ [NSP2] or 5’-GTCCAATTCAT-NCCTGGTG-3’ [VP6]), 0.25 μl of 10 μM probe (FAM5’-TCCAATAGATTGAAGTCAGTAACGT-TTCCA-3’MGB [NSP2] or FAM5’-CCACCRAAYATGACRCCAGCNGTA-3’MGB [VP6]), and 6.25 μl of nuclease-free water (ThermoFisher Scientific). In a 96-well plate, 23 μl of master mix was combined with 2 μl of cDNA. Six standards for the target gene as well as a no-template control and a positive control were added to each PCR plate. The plate was sealed with a Microseal ‘C’ Film (Bio-Rad) and subjected to the following cycles in a Bio-Rad CFX qPCR Instrument (Bio-Rad): initial UDG-incubation 50°C for 2 minutes; denaturation for 95°C for 2 minutes; then 40 cycles of 95°C for 15 seconds and 60°C for 1 minute. Data were considered valid if amplification was absent in the negative control and present in the positive control. qPCR standards consisting of TOPO-TA plasmid constructs containing either NPS2 or VP6 gene amplicons (genotype 1) were produced at the University of Liverpool and distributed to the other study sites.

### Inflammatory biomarkers

For infant stools collected before each dose of ORV, MPO and α1AT concentrations were measured by ELISA at each site. ELISA kits with the same lot number were distributed across the study sites. Stool samples were defrosted on ice and 100 mg then aliquoted into a polypropylene tube. ELISAs were carried out following the manufacturer’s instructions (IDK 2018 instructions for MPO and BioVendor RIC6200 for α1AT). Paired samples from a given infant were run on the same plate, and MPO and α1AT assays were carried out in tandem to avoid freeze-thaw cycles. For week 6 serum samples of Indian and Malawian infants, α1AG was measured at CMC, Vellore, using a commercial ELISA kit (abcam) according to the manufacturer’s instructions.

### DNA extraction from stool

DNA extraction from stool was carried out using the QIAamp DNA stool mini kit (Qiagen) according to the manufacturer’s instructions (QIAamp DNA Stool Handbook 06/2012), with several modifications to increase DNA yield. Briefly, the vortexed suspension of ASL buffer and stool was added to 370 mg of 0.1 mm zirconia-silicate beads (BioSpec) with 1.67 μl of lyzozyme (30 mg/ml) (Sigma-Aldrich). Samples were then incubated at 37°C on a shaker (Eppendorf Thermo Mixer Comfort) at 250-300 rpm for 10 minutes. 10 μl of proteinase K (Qiagen), 50 μl of 10% sodium dodecyl sulphate (Sigma-Aldrich), and 20 μl of RNase A (1mg/ml; Sigma-Aldrich) was added before incubation on a heating block (Eppendorf Thermo Mixer Comfort) at 70°C for 10 minutes. After allowing the samples to cool for 3 minutes, bead beating was performed for 5 minutes using a Tissue Lyser II (Qiagen) at 25 Hz. Samples were microcentrifuged at 16,100 g for 1 minute and the supernatant was then transferred into a new microcentrifuge tube containing an InhibitEX tablet and vortexed for 1 minute. The suspension was incubated at room temperature for 1 minute, centrifuged at 16,100 g for 5 minutes, and the supernatant (approximately 600 μl) then transferred into a new 2 ml tube. The supernatant was centrifuged at 16,100 g for 3 minutes, then 400 μl added to a 2 ml tube containing 400 μl of buffer AL. 400 μl of ethanol (96–100%) was added to the lysate and mixed by vortexing. The manufacturer’s instructions were then resumed until elution in 50 μl of AE Buffer. An extraction control was carried out with each batch of DNA extraction at all sites. Extracted DNA was shipped from India and Malawi to the University of Liverpool on dry ice for 16S rRNA sequencing.

### Amplicon generation for 16S rRNA sequencing

Amplicons spanning 16S rRNA gene variable regions 3 and 4 (primers 309F 5’-overhang-ACTCCTACGGGAGGCAGCAG-3’ and 819R 5’-overhang-GGACTACHVGGGTWTCTAAT-3’) were produced following the established Illumina protocol (Illumina, 16S Metagenomic Sequencing Library Preparation Protocol Part # 15044223 Rev. B) with the following amendments: 1 μl of DNA was used as a starting template; NEBNext Q5 Hot Start HiFi PCR Master Mix (NEB) was used for amplicon and indexing PCRs; PCR reaction volumes were halved to 25 μl; primers were used at a concentration of 10 μM; and 20 μg of molecular-grade bovine serum albumin (NEB) was added to the amplicon PCR master mix to mitigate the effect of PCR inhibitors. Cycling conditions for amplicon PCR involved: denaturation at 98°C for 30 seconds; 10 cycles of 98°C for 10 seconds, 55°C for 15 seconds, and 72°C for 40 seconds; and final extension at 72°C for 60 seconds. Cycling conditions for indexing PCR involved: denaturation at 98°C for 3 minutes; 15 cycles of 98°C for 10 seconds, 55°C for 15 seconds, and 72°C for 40 seconds; and final extension at 72°C for 5 minutes. AMPure XP beads were substituted with a custom preparation of Sera-Mag SpeedBeads Protein A/G particles (MERCK/GE Healthcare) throughout the protocol. To determine volumes required for equimolar pooling, amplicon concentrations were determined by Quanti-it (ThermoFisher Scientific) and size distributions by fragment analysis on a 5300 Fragment Analyzer System (Agilent). Amplicons were pooled using a mosquito X1 (TTP Labtech) liquid handling robot. The final library underwent size selection to remove potentially contaminating primer-dimers and genomic DNA using the Pippin Prep (Sage Science) 1.5% Agarose Gel Cassette (Labtech).

Overall, we sequenced amplicons from 2,138 samples across 14 Illumina HiSeq2500 lanes (v2 chemistry, 600 cycles in rapid run mode) and one Illumina MiSeq lane (v3 chemistry, 600 cycles). Since enrolment and sample collection across the three sites were not synchronised, sequencing was batched by geographic origin according to sample availability. Samples from a mother–infant pair were processed on the same PCR plate. Each PCR plate contained a no-template PCR control, stool controls provided by a mother and infant in the UK who were not enrolled in the study, DNA from a mock community (Zymo Research D6306), and a pool of extraction controls corresponding to the samples contained on the PCR plate. Final libraries consisted of up to four PCR plates (384 amplicons). Eight samples were excluded from the analysis owing to the presence of significant amplification from their corresponding extraction controls (**Figure S8**).

### Illumina sequencing of 16S rRNA libraries

The quantity and quality of each amplicon pool was assessed by Qubit HS DNA kit (ThermoFisher Scientific), Bioanalyzer High Sensitivity DNA Kit (Agilent), and qPCR using the KAPA Library Quantification Kit (Roche) on a Roche Light Cycler LC480II (Roche) according to the manufacturer’s instructions. qPCR data were used to calculate sample molarity. 5 μl of the final pool was denatured for 5 minutes at room temperature using 5 μl of freshly diluted 0.1 N sodium hydroxide (Illumina). The reaction was subsequently terminated by the addition of HT1 hybridization buffer (Illumina) and the library diluted post-denaturation to a final loading concentration of 7–8.5 pM. Libraries were sequenced with 10–20% PhiX (Illumina) using the 2×300 bp paired-end protocol. The raw sequence data generated during this study have been deposited in the European Nucleotide Archive (accession number PRJEB38948).

### Independent validation of 16S rRNA sequencing

To validate the robustness of the microbiota protocol, 10% of DNA samples were shipped to Imperial College London and sequenced according to the methods described above. We included 30 mother–infant pairs per study site in the validation subset and sequenced two infant stools (from weeks 1 and 10/12) and the maternal stool from each pair. Protocol deviations included the use of AMPure XP beads for PCR product purification, fragment analysis via TapeStataion D1000 ScreenTape (Agilent Technologies), concentration quantification via Qubit dsDNA High Sensitivity Assay Kit (ThermoFisher Scientific), and manual pooling of equimolar quantities. The order of samples was randomised across three PCR plates and the pooled libraries were sequenced with 10–20% PhiX (Illumina) on two 2×300 bp paired-end Illumina MiSeq runs (v3 chemistry).

### Bioinformatic processing of sequence data

Adapters were trimmed from raw sequences using cutadapt version 1.18 ^38^. Subsequent steps for quality filtering, denoising, merging of paired reads, and chimera removal followed the DADA2 pipeline ^39^ implemented in QIIME 2 with default parameters ^40^. Forward reads were truncated to 270 bp and reverse reads to 200 bp to account for the fall in sequencing quality scores towards the end of each read. Sequence data were processed separately for each run, and the feature tables and corresponding sequences then merged. All subsequent steps were performed in the programming language R. Taxonomy assignment was performed using the RDP naïve Bayesian classifier trained on the Silva rRNA database (version 132). Sequence variants were included in the analysis if they were 390–440 bp in length, bacterial, detectable at an abundance of ≥0.1% in at least two samples, and passed frequency-based contamination filtering using the *decontam* package (which screens sequences that are more abundant in low-concentration samples) ^41^. Nanodrop concentrations (ng/μl) of extracted DNA served as the basis for contaminant filtering. Samples with at least 25,000 quality-filtered sequences were retained in the final analysis.

### Statistical analysis

Processed data and analysis code generated during this study are available on github (https://github.com/eparker12/RoVI). All analyses were performed in the programming language R. Seroconversion and shedding proportions were compared by country using pairwise Fisher’s exact tests with Benjamini–Hochberg FDR correction. Antibody and inflammatory biomarker concentrations were log transformed and compared by country and shedding subgroup (see **Figure 1E**) using ANOVA with post-hoc Tukey tests. To explore covariates associated with ORV outcome, we used either logistic regression (for seroconversion and shedding status) or linear regression and Pearson’s *r* (for log-transformed post-vaccination RV-IgA). In Indian mother–infant dyads, Spearman’s rank correlation coefficients were calculated between all measured antibody concentrations (RV-IgG and RV-IgA) and rotavirus shedding quantities (reciprocal of qPCR Ct value). A hierarchical clustering analysis of all infant and maternal gut microbiota samples was conducted using Ward’s minimum variance hierarchical clustering method based on the presence or absence of common genera (≥1% prevalence at ≥0.1% abundance). Longitudinal mixed-effects models were used to compare microbiota diversity (Shannon index, genus-level richness) by country (pairwise comparisons with FDR correction) and ORV outcome (seroconversion and shedding status), including week as a covariate and study ID as a random effect. We used longitudinal zero-inflated negative binomial models of genus counts to identify taxa that discriminated infants according to country, seroconversion, and shedding status, adjusting for age (by including week as a fixed effect) and including study ID as a random effect. For taxa with a prevalence of >95%, negative binomial models without zero inflation were used. Fisher’s exact test was also used for cross-sectional comparisons of genus prevalence (≥0.1% abundance threshold) by country, excluding taxa that were ubiquitous (>95% in both comparison groups) or scarce (<5% in both groups). We report on genera with FDR-adjusted p values of <0.1.

We applied the Random Forests algorithm in a series of cross-sectional analyses to predict country, seroconversion, or shedding status (classification approach), or post-vaccination log-transformed RV-IgA concentration (regression approach) based on genus relative abundances. For each analysis, we performed 20 iterations of 5-fold cross-validation. For classification models (of binary outcomes), we standardised the baseline accuracy at 50% by fitting each iteration of 5-fold cross-validation on a random subset of 50 samples per group (or the number of samples in the minority group if this was <50). Models were excluded if there were <10 samples in the minority group. Mean cross-validation variable importance was determined based on the increase in Gini index (classification) or mean squared error (regression) of out-of-bag sample prediction following random permutation. For Indian infants, we assessed additional models based on demography/baseline health measurements (18 variables), exposure/antibody data (12 variables, encompassing inflammatory biomarkers, wild-type rotavirus shedding, and maternal antibodies), and genus relative abundances (126 variables), and all variables combined.

For technical replicates (including positive controls and samples selected for independent validation), we quantified the proportion of variance explained by sample ID using linear regression (alpha diversity, genus abundances) or PERMANOVA with 999 permutations (weighted and unweighted Bray–Curtis distances). We also used PERMANOVA to quantify the proportion of variation in microbiota composition associated with sequencing run, stratified by age and country.

## Supplemental table titles and legends

**Table S1. Sample counts for primary outcomes**. α1AG, α1 acid glycoprotein; α1AT, α1-antitrypsin; MPO, myeloperoxidase; neo+, infected with rotavirus pre-vaccination; neo-, uninfected with rotavirus pre-vaccination; RV, rotavirus.

**Table S2. Cofactors associated with neonatal rotavirus infection**. α1AG, α1 acid glycoprotein; α1AT, α1-antitrypsin; bOPV, bivalent oral poliovirus vaccine; GMC, geometric mean concentration; IPV, inactivated poliovirus vaccine; LRT, likelihood ratio test for fit of model with vs without polio vaccine schedule; MPO, myeloperoxidase; ORV, oral rotavirus vaccine; RR, relative risk; RV, rotavirus; tOPV, trivalent oral poliovirus vaccine; *log-transformed to approximate normality in statistical models.

**Table S3A. Cofactors associated with rotavirus vaccine shedding**. Data are presented for (A) dose 1 shedding and (B) shedding after either dose. Measurements obtained after the first dose of ORV are excluded to focus on potential predictors of ORV shedding rather than shedding–induced changes. α1AG, α1 acid glycoprotein; α1AT, α1–antitrypsin; bOPV, bivalent oral poliovirus vaccine; GMC, geometric mean concentration; IPV, inactivated poliovirus vaccine; LRT, likelihood ratio test for fit of model with vs without polio vaccine schedule; MPO, myeloperoxidase; ORV, oral rotavirus vaccine; RR, relative risk; RV, rotavirus; tOPV, trivalent oral poliovirus vaccine; *log–transformed to approximate normality in statistical models; † excluded to minimise multicollinearity.

**Table S4. Cofactors associated with rotavirus seroconversion**. α1AG, α1 acid glycoprotein; α1AT, α1-antitrypsin; bOPV, bivalent oral poliovirus vaccine; GMC, geometric mean concentration; IPV, inactivated poliovirus vaccine; LRT, likelihood ratio test for fit of model with vs without polio vaccine schedule; MPO, myeloperoxidase; ORV, oral rotavirus vaccine; RR, relative risk; RV, rotavirus; tOPV, trivalent oral poliovirus vaccine; *log-transformed to approximate normality in statistical models; † excluded to minimise multicollinearity.

**Table S5. Cofactors associated with post-vaccination RV-IgA concentration**. α1AG, α1 acid glycoprotein; α1AT, α1-antitrypsin; bOPV, bivalent oral poliovirus vaccine; GMC, geometric mean concentration; IPV, inactivated poliovirus vaccine; LRT, likelihood ratio test for fit of model with vs without polio vaccine schedule; MPO, myeloperoxidase; ORV, oral rotavirus vaccine; RV, rotavirus; tOPV, trivalent oral poliovirus vaccine; *log-transformed to approximate normality in statistical models; † excluded to minimise multicollinearity.

**Table S6. Geographic discrepancies in genus prevalence**. Data are presented for (A) Malawi vs India, (B) India vs UK, and (C) Malawi vs UK. Genus prevalences were compared using Fisher’s exact test, with a 0.1% abundance threshold to define presence/absence. Taxa were excluded if ubiquitous (>95% in both groups) or scarce (<5% in both groups).

**Table S7. Taxa correlated with ORV response in longitudinal zero-inflated negative binomial models**. FDR, false discovery rate; ORV, oral rotavirus vaccine; RV, rotavirus.

## Notes

### Clinical Trial

CTRI/2015/11/006354

### Clinical Protocols

https://bmjopen.bmj.com/content/7/3/e016577.long

### Author Declarations

The study was approved by the Institutional Review Board at the Christian Medical College (CMC) in Vellore, the College of Medicine Research and Ethics Committee in Blantyre, and the North West - Liverpool East Research Ethics Committee in Liverpool.

