## Supplementary Figures S1-S8 for "Impact of maternal antibodies and microbiota development on the immunogenicity of oral rotavirus vaccine in African, Indian, and European infants: a prospective cohort study"

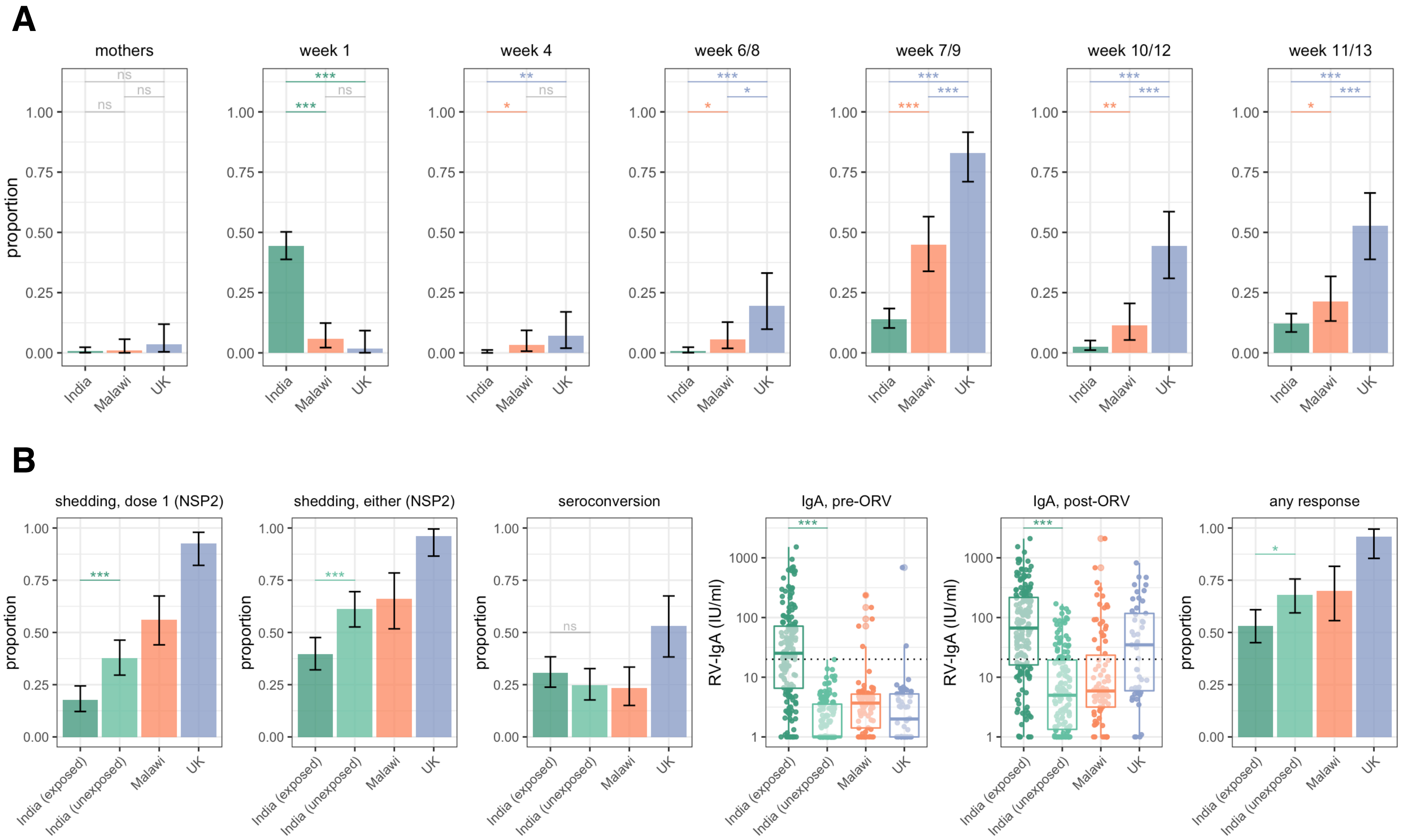
A
mothers
week 1
week 4
week 6/8
week 7/9
week 10/12
week 11/13
proportion
India
Malawi
UK
B
shedding, dose 1 (NSP2)
shedding, either (NSP2)
seroconversion
IgA, pre-ORV
IgA, post-ORV
any response
RV-IgA (IU/ml)
India (exposed)
India (unexposed)

Figure S1

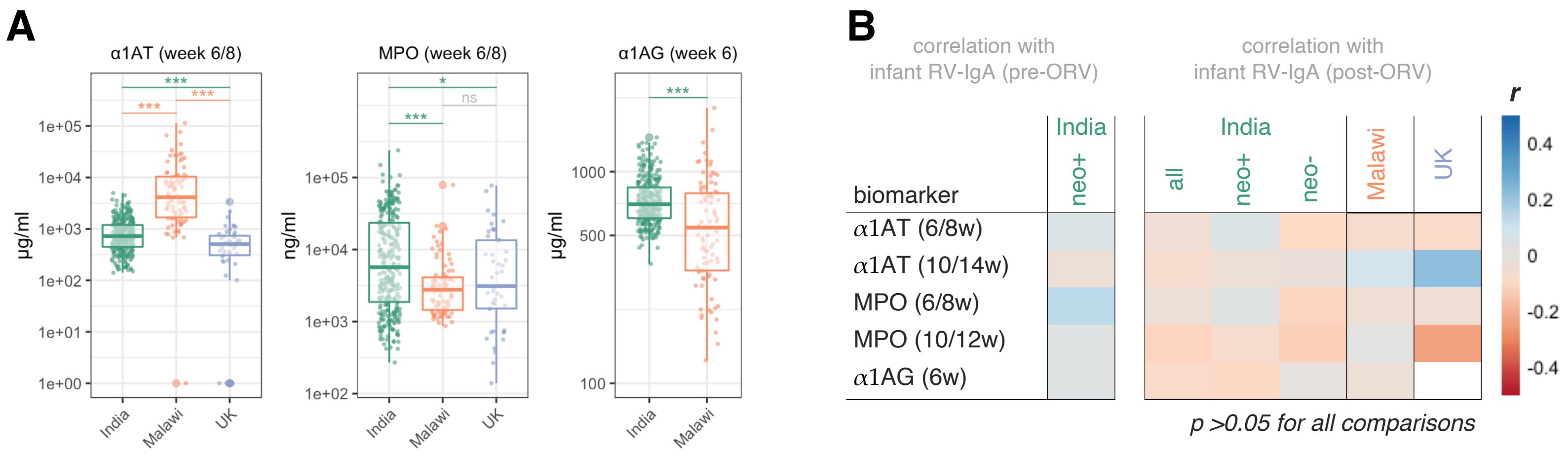
A
α1AT (week 6/8)
MPO (week 6/8)
α1AG (week 6)
µg/ml
ng/ml
India
Malawi
UK
***
ns
*
B
correlation with
infant RV-IgA (pre-ORV)
correlation with
infant RV-IgA (post-ORV)
biomarker
India
neo+
all
neo-
Malawi
UK
α1AT (6/8w)
α1AT (10/14w)
MPO (6/8w)
MPO (10/12w)
α1AG (6w)
r
p >0.05 for all comparisons

Figure S2

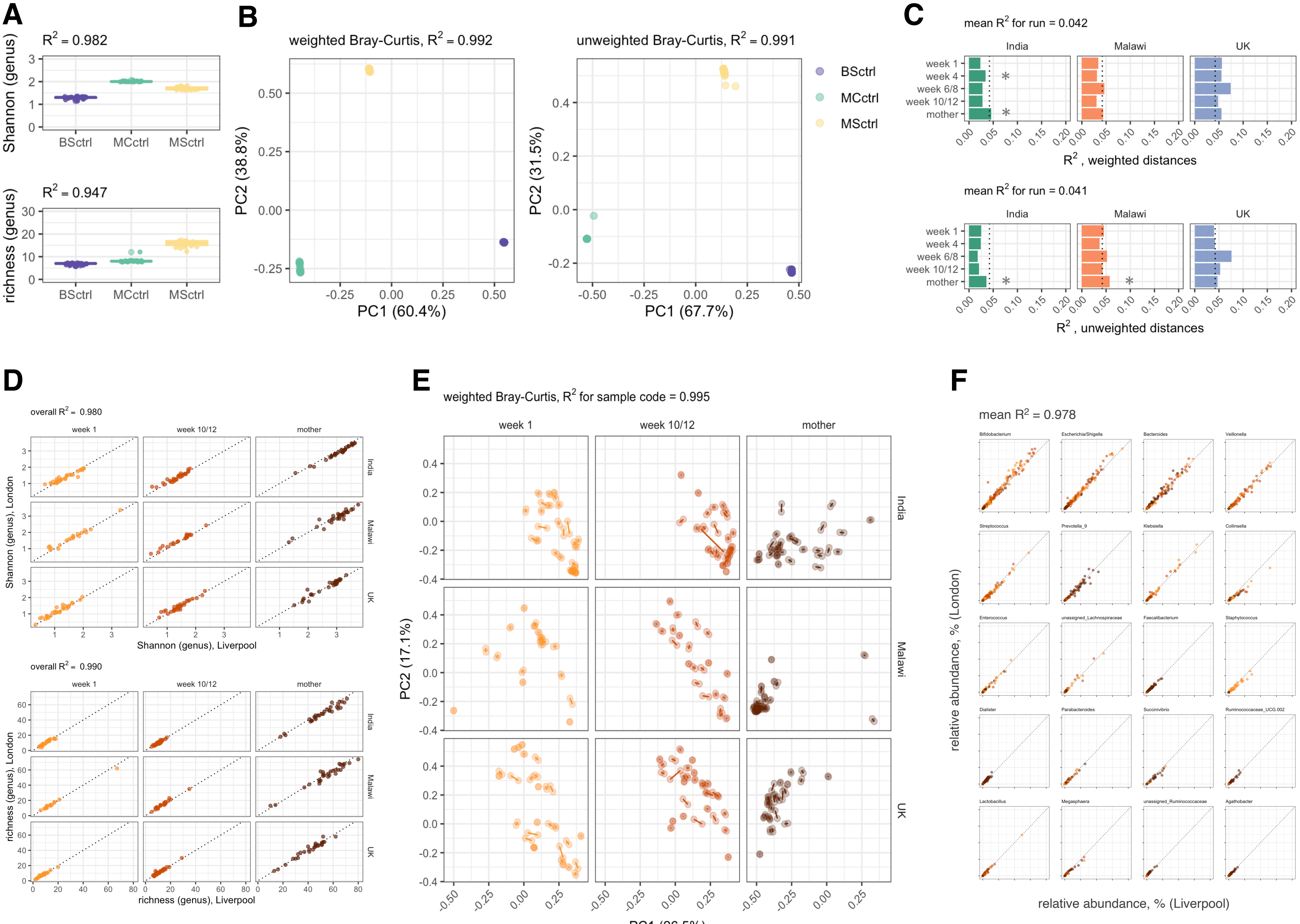

A
$R^2$ = 0.982
Shannon (genus)
BSctrl
MCctrl
MSctrl
$R^2$ = 0.947
richness (genus)
B
weighted Bray-Curtis, $R^2$ = 0.992
unweighted Bray-Curtis, $R^2$ = 0.991
PC2 (38.8%)
PC1 (60.4%)
PC2 (31.5%)
PC1 (67.7%)
C
mean $R^2$ for run = 0.042
India
Malawi
UK
week 1
week 4
week 6/8
week 10/12
mother
$R^2$ , weighted distances
mean $R^2$ for run = 0.041
$R^2$ , unweighted distances
D
overall $R^2$ = 0.980
Shannon (genus), London
Shannon (genus), Liverpool
overall $R^2$ = 0.990
richness (genus), London
richness (genus), Liverpool
E
weighted Bray-Curtis, $R^2$ for sample code = 0.995
PC2 (17.1%)
PC1 (26.5%)
F
mean $R^2$ = 0.978
Bifidobacterium
Escherichia/Shigella
Bacteroides
Veillonella
Streptococcus
Prevotella_9
Klebsiella
Collinsella
Enterococcus
unassigned_Lachnospiraceae
Faecalibacterium
Staphylococcus
Dialister
Parabacteroides
Succinivibrio
Ruminococcaceae_UCG.002
Lactobacillus
Megasphaera
unassigned_Ruminococcaceae
Agathobacter
relative abundance, % (London)
relative abundance, % (Liverpool)


Figure S3

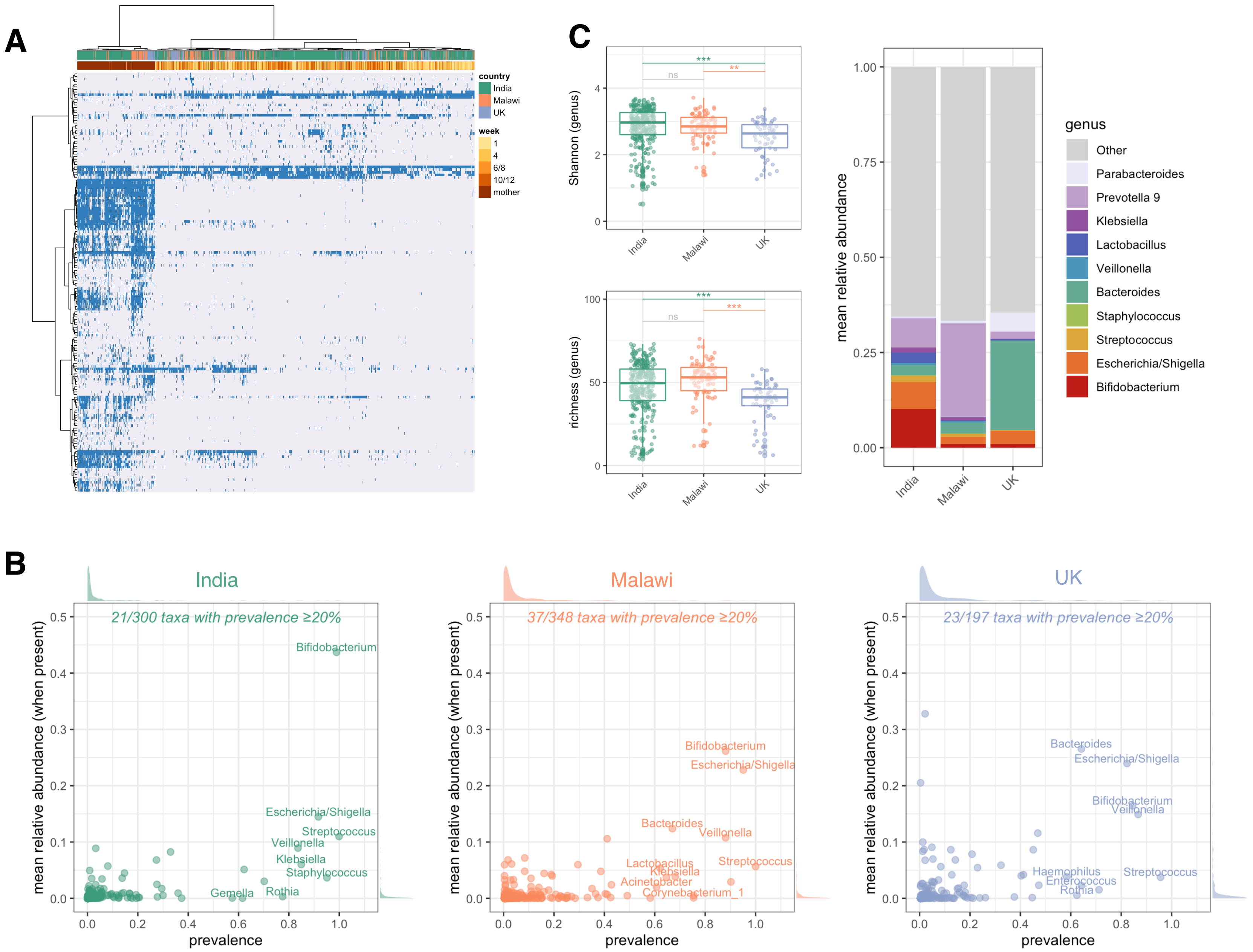
A
country
India
Malawi
UK
week
1
4
6/8
10/12
mother
C
Shannon (genus)
richness (genus)
***
**
ns
India
Malawi
UK
mean relative abundance
genus
Other
Parabacteroides
Prevotella 9
Klebsiella
Lactobacillus
Veillonella
Bacteroides
Staphylococcus
Streptococcus
Escherichia/Shigella
Bifidobacterium
B
India
21/300 taxa with prevalence ≥20%
Malawi
37/348 taxa with prevalence ≥20%
UK
23/197 taxa with prevalence ≥20%
mean relative abundance (when present)
prevalence
Bifidobacterium
Escherichia/Shigella
Streptococcus
Veillonella
Klebsiella
Staphylococcus
Gemella
Rothia
Bacteroides
Lactobacillus
Acinetobacter
Corynebacterium_1
Haemophilus
Enterococcus

Figure S4

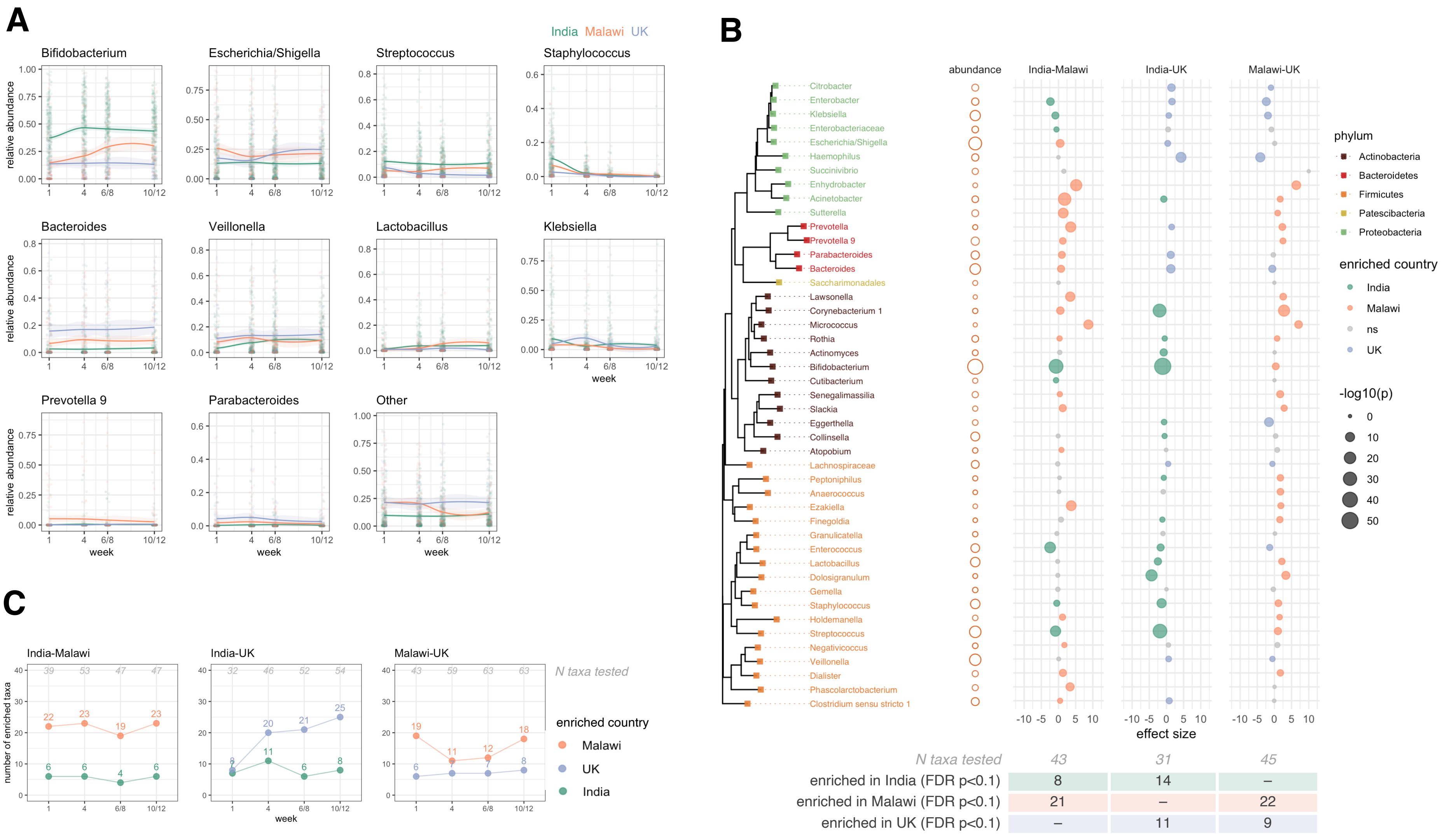

A
India Malawi UK
Bifidobacterium
Escherichia/Shigella
Streptococcus
Staphylococcus
Bacteroides
Veillonella
Lactobacillus
Klebsiella
Prevotella 9
Parabacteroides
Other
relative abundance
week
B
abundance
India-Malawi
India-UK
Malawi-UK
Citrobacter
Enterobacter
Klebsiella
Enterobacteriaceae
Escherichia/Shigella
Haemophilus
Succinivibrio
Enhydrobacter
Acinetobacter
Sutterella
Prevotella
Prevotella 9
Parabacteroides
Bacteroides
Saccharimonadales
Lawsonella
Corynebacterium 1
Micrococcus
Rothia
Actinomyces
Bifidobacterium
Cutibacterium
Senegalimassilia
Slackia
Eggerthella
Collinsella
Atopobium
Lachnospiraceae
Peptoniphilus
Anaerococcus
Ezakiella
Finegoldia
Granulicatella
Enterococcus
Lactobacillus
Dolosigranulum
Gemella
Staphylococcus
Holdemanella
Streptococcus
Negativicoccus
Veillonella
Dialister
Phascolarctobacterium
Clostridium sensu stricto 1
effect size
phylum
Actinobacteria
Bacteroidetes
Firmicutes
Patescibacteria
Proteobacteria
enriched country
India
Malawi
ns
UK
-log10(p)
N taxa tested 43 31 45
enriched in India (FDR p<0.1) 8 14 –
enriched in Malawi (FDR p<0.1) 21 – 22
enriched in UK (FDR p<0.1) – 11 9
C
India-Malawi
India-UK
Malawi-UK
N taxa tested
enriched country
Malawi
UK
India
number of enriched taxa
week


Figure S5

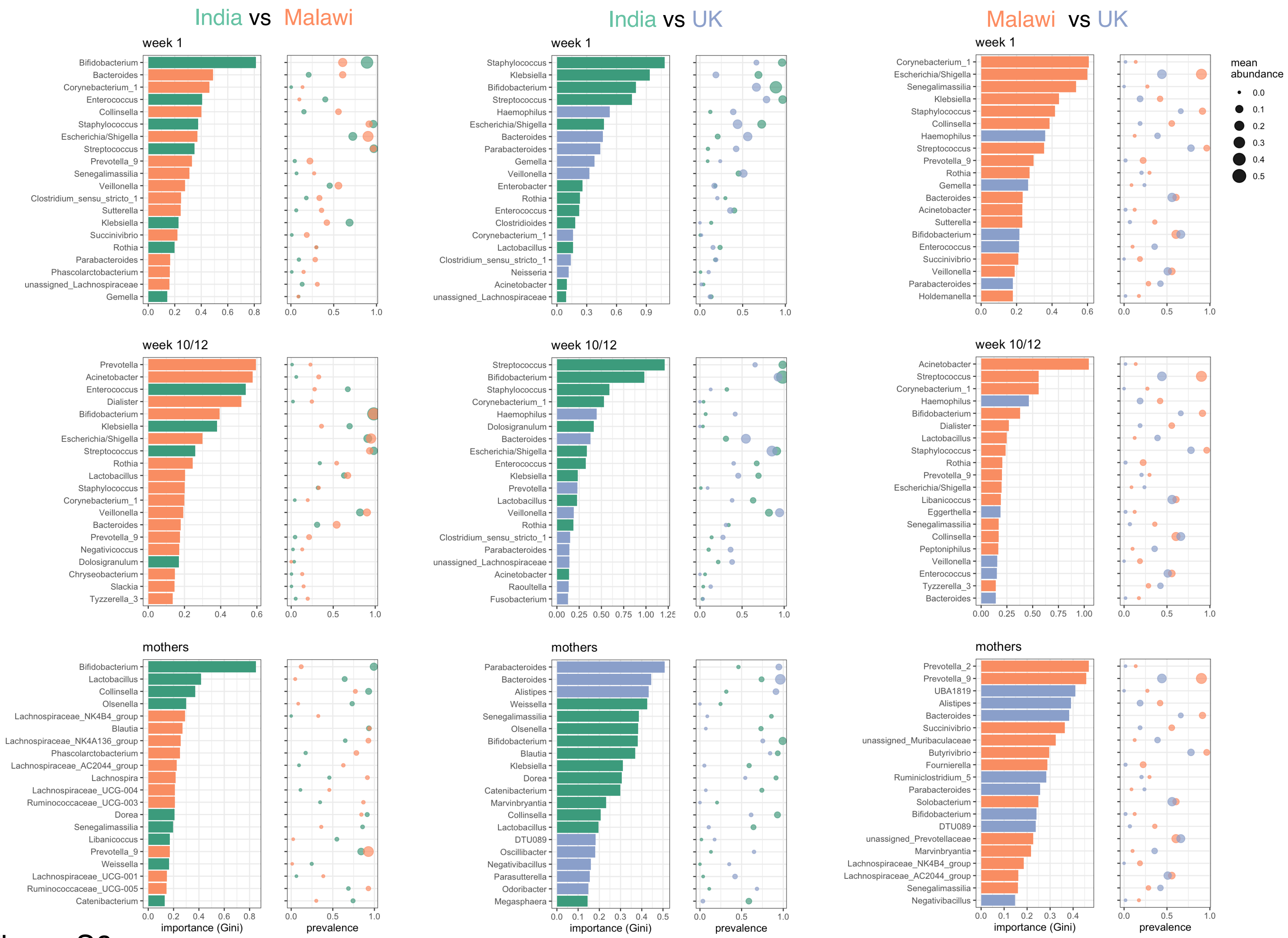

India vs Malawi
India vs UK
Malawi vs UK
week 1
Bifidobacterium
Bacteroides
Corynebacterium_1
Enterococcus
Collinsella
Staphylococcus
Escherichia/Shigella
Streptococcus
Prevotella_9
Senegalimassilia
Veillonella
Clostridium_sensu_stricto_1
Sutterella
Klebsiella
Succinivibrio
Rothia
Parabacteroides
Phascolarctobacterium
unassigned_Lachnospiraceae
Gemella
week 1
Staphylococcus
Klebsiella
Bifidobacterium
Streptococcus
Haemophilus
Escherichia/Shigella
Bacteroides
Parabacteroides
Gemella
Veillonella
Enterobacter
Rothia
Enterococcus
Clostridioides
Corynebacterium_1
Lactobacillus
Clostridium_sensu_stricto_1
Neisseria
Acinetobacter
unassigned_Lachnospiraceae
week 1
Corynebacterium_1
Escherichia/Shigella
Senegalimassilia
Klebsiella
Staphylococcus
Collinsella
Haemophilus
Streptococcus
Prevotella_9
Rothia
Gemella
Bacteroides
Acinetobacter
Sutterella
Bifidobacterium
Enterococcus
Succinivibrio
Veillonella
Parabacteroides
Holdemanella
mean abundance
week 10/12
Prevotella
Acinetobacter
Enterococcus
Dialister
Bifidobacterium
Klebsiella
Escherichia/Shigella
Streptococcus
Rothia
Lactobacillus
Staphylococcus
Corynebacterium_1
Veillonella
Bacteroides
Prevotella_9
Negativicoccus
Dolosigranulum
Chryseobacterium
Slackia
Tyzzerella_3
week 10/12
Streptococcus
Bifidobacterium
Staphylococcus
Corynebacterium_1
Haemophilus
Dolosigranulum
Bacteroides
Escherichia/Shigella
Enterococcus
Klebsiella
Prevotella
Lactobacillus
Veillonella
Rothia
Clostridium_sensu_stricto_1
Parabacteroides
unassigned_Lachnospiraceae
Acinetobacter
Raoultella
Fusobacterium
week 10/12
Acinetobacter
Streptococcus
Corynebacterium_1
Haemophilus
Bifidobacterium
Dialister
Lactobacillus
Staphylococcus
Rothia
Prevotella_9
Escherichia/Shigella
Libanicoccus
Eggerthella
Senegalimassilia
Collinsella
Peptoniphilus
Veillonella
Enterococcus
Tyzzerella_3
Bacteroides
mothers
Bifidobacterium
Lactobacillus
Collinsella
Olsenella
Lachnospiraceae_NK4B4_group
Blautia
Lachnospiraceae_NK4A136_group
Phascolarctobacterium
Lachnospiraceae_AC2044_group
Lachnospira
Lachnospiraceae_UCG-004
Ruminococcaceae_UCG-003
Dorea
Senegalimassilia
Libanicoccus
Prevotella_9
Weissella
Lachnospiraceae_UCG-001
Ruminococcaceae_UCG-005
Catenibacterium
mothers
Parabacteroides
Bacteroides
Alistipes
Weissella
Senegalimassilia
Olsenella
Bifidobacterium
Blautia
Klebsiella
Dorea
Catenibacterium
Marvinbryantia
Collinsella
Lactobacillus
DTU089
Oscillibacter
Negativibacillus
Parasutterella
Odoribacter
Megasphaera
mothers
Prevotella_2
Prevotella_9
UBA1819
Alistipes
Bacteroides
Succinivibrio
unassigned_Muribaculaceae
Butyrivibrio
Fournierella
Ruminiclostridium_5
Parabacteroides
Solobacterium
Bifidobacterium
DTU089
unassigned_Prevotellaceae
Marvinbryantia
Lachnospiraceae_NK4B4_group
Lachnospiraceae_AC2044_group
Senegalimassilia
Negativibacillus
importance (Gini)
prevalence
importance (Gini)
prevalence
importance (Gini)
prevalence


Figure S6

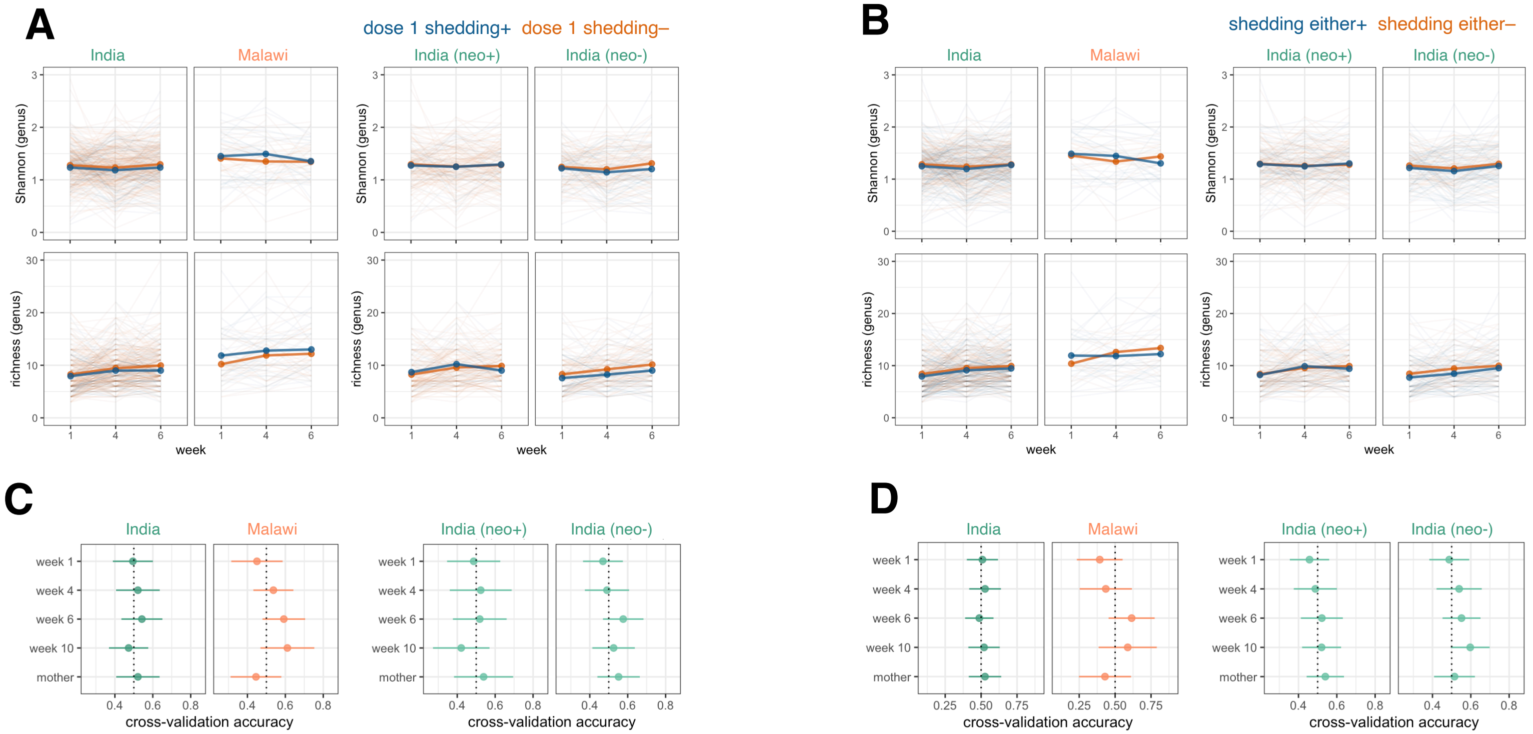


**E** longitudinal ZINB models of taxon abundance

| | India all | India neo+ | India neo- | Malawi | UK |
|---|---|---|---|---|---|
| N genera tested* | 23 | 24 | 22 | 43 | nf |
| + correlation with shedding (dose 1) | 0 | 0 | 0 | 0 | nf |
| – correlation with shedding (dose 1) | 0 | 0 | 0 | 1 | nf |
| N genera tested* | 23 | 24 | 22 | 43 | nf |
| + correlation with shedding (either) | 0 | 0 | 0 | 4 | nf |
| – correlation with shedding (either) | 2 | 0 | 1 | 2 | nf |

**F**

post-ORV RV-IgA

| | India all | India neo+ | India neo- | Malawi | UK |
|---|---|---|---|---|---|
| week 1 | 287 | 155 | 131 | 73 | 53 |
| week 4 | 285 | 156 | 128 | 71 | 53 |
| week 6/8 | 287 | 154 | 132 | 66 | 48 |
| week 10/12 | 281 | 160 | 120 | 53 | 51 |
| maternal | 286 | 156 | 129 | 74 | 51 |

seroconversion

| | India all | India neo+ | India neo- | Malawi | UK |
|---|---|---|---|---|---|
| week 1 | 100 | 94 | 64 | 36 | 46 |
| week 4 | 100 | 92 | 60 | 30 | 48 |
| week 6/8 | 100 | 92 | 66 | 24 | 46 |
| week 10/12 | 100 | 98 | 60 | nf | 48 |
| maternal | 100 | 92 | 62 | 32 | 46 |

shedding, dose 1

| | India all | India neo+ | India neo- | Malawi | UK |
|---|---|---|---|---|---|
| week 1 | 100 | 52 | 92 | 58 | nf |
| week 4 | 100 | 56 | 96 | 64 | nf |
| week 6/8 | 100 | 52 | 98 | 50 | nf |
| week 10/12 | – | – | – | – | – |
| maternal | 100 | 52 | 92 | 54 | nf |

shedding, either dose

| | India all | India neo+ | India neo- | Malawi | UK |
|---|---|---|---|---|---|
| week 1 | 100 | 100 | 100 | 32 | nf |
| week 4 | 100 | 100 | 98 | 30 | nf |
| week 6/8 | 100 | 100 | 100 | 32 | nf |
| week 10/12 | – | – | – | – | – |
| maternal | 100 | 100 | 100 | 28 | nf |

Figure S7

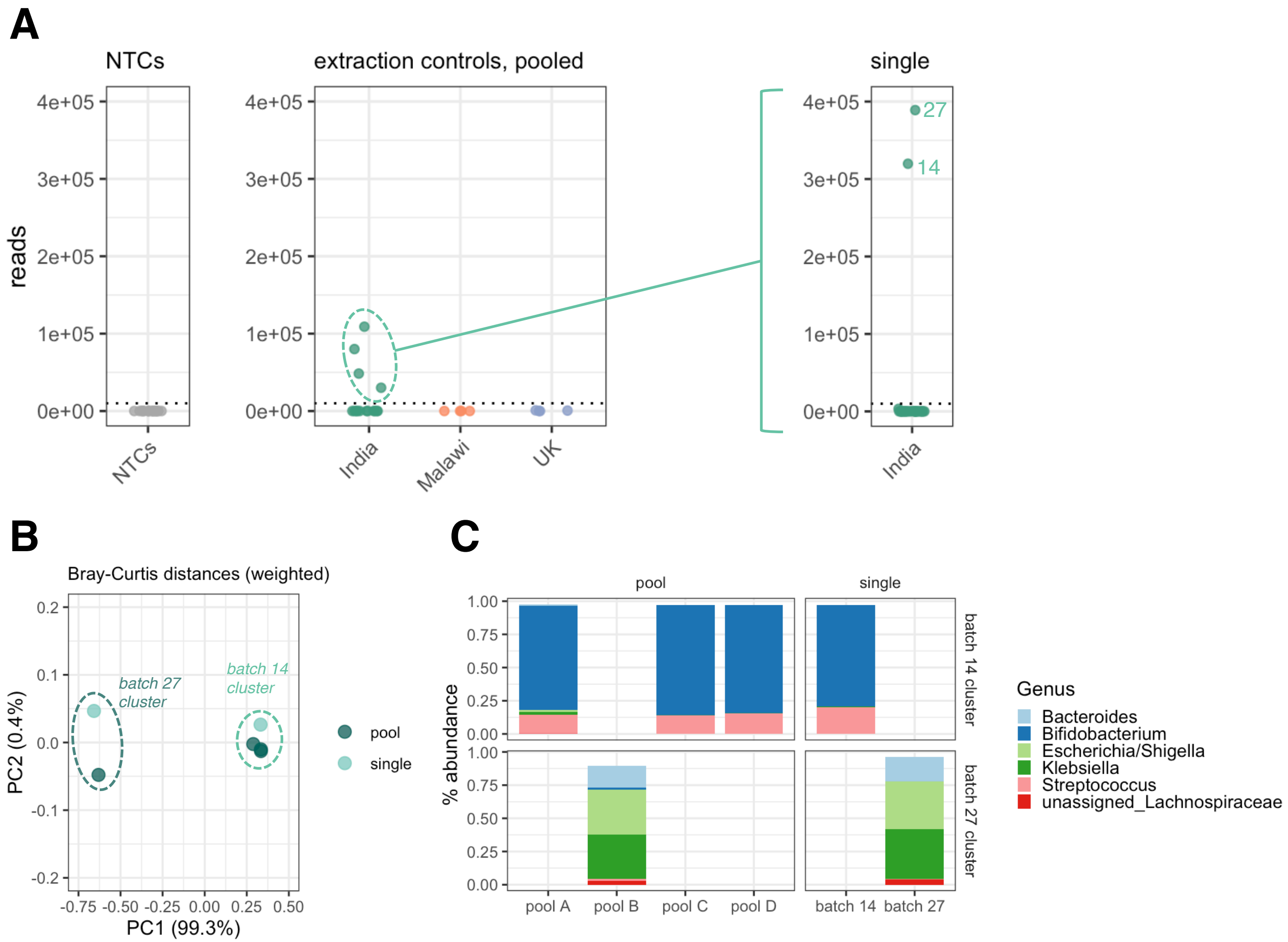
A
NTCs
extraction controls, pooled
single
reads
4e+05
3e+05
2e+05
1e+05
0e+00
NTCs
India
Malawi
UK
27
14
India
B
Bray-Curtis distances (weighted)
batch 27
cluster
batch 14
cluster
PC2 (0.4%)
PC1 (99.3%)
pool
single
C
pool
single
% abundance
batch 14 cluster
batch 27 cluster
pool A
pool B
pool C
pool D
batch 14
batch 27
Genus
Bacteroides
Bifidobacterium
Escherichia/Shigella
Klebsiella
Streptococcus
unassigned_Lachnospiraceae

Figure S8
